# Analysis of Alzheimer’s disease Polygenic Risk Scores using RNA-sequencing provides further novel biological pathways

**DOI:** 10.1101/2022.06.29.22276952

**Authors:** K. Crawford, G. Leonenko, E. Baker, D. Grozeva, B. Lan-Leung, P. Holmans, J. Williams, M. C. O’Donovan, V. Escott-Price, DK. Ivanov

**Affiliations:** UK Dementia Research Institute at Cardiff (UKDRI), Cardiff University, College of Biomedical and Life Sciences, Hadyn Ellis Building, Cardiff CF24 4HQ, UK; Centre for Trials Research, Cardiff University, Cardiff CF24 4HQ, UK; MRC Centre for Neuropsychiatric Genetics and Genomics, Division of Psychological Medicine and Clinical Neurosciences, Cardiff University, School of Medicine, Hadyn Ellis Building, Cardiff CF24 4HQ, UK

## Abstract

Polygenic risk scores (PRS) have been widely adopted as a tool for measuring common variant liability and it has been shown to predict lifetime risk of Alzheimer’s disease (AD) development. However, the relationship between PRS and AD pathogenesis is largely unknown. We aimed to address some of the knowledge gaps with respect to the downstream molecular consequences associated with PRS. We also make a direct comparison of the disrupted biological mechanisms in a case/control classification and in response to PRS in the same individuals.

We performed an integrative computational analysis of the transcriptome of the largest human brain-derived cohort sample (288 individuals; cerebellum and temporal cortex; MayoRNAseq; AMP-AD) with matched AD genetic and gene-expression data (WGS; bulk-brain RNA-seq). There was little overlap in terms of differentially expressed genes in case/control and PRS analyses, but a consensus of commonly disrupted biological mechanisms. Genes implicated by previous AD GWAS were found to be significantly enriched with respect to PRS in temporal cortex only. We identified mechanisms that were previously implicated in AD, including immune/stress response, lipid/cholesterol/fatty acid metabolism, endosome, death/apoptosis, neuronal processes, ageing and the involvement of glial cells. We also provide novel evidence for the significant involvement in AD of cellular structures, including the Golgi apparatus and endoplasmic reticulum as well as mitochondrial function.

The largely common biological mechanisms between a case/control classification and in association with PRS suggests that PRS stratification can be used for studies where suitable case/control samples are not available or the selection of individuals with high and low PRS in clinical trials.

## 1. Introduction

Alzheimer’s disease (AD) is a neurodegenerative disorder characterised by progressive cognitive decline, molecular changes including, but not limited to the accumulation of beta-amyloids (extracellular Aß plaques) and tau tangles in the human brain^1^. The molecular changes are detectable much earlier than the clinical phenotype, occurring for example 10-20 years before cognitive deterioration^2^. Currently, there are no approved pharmacological or other treatments that have been shown to reverse or stop the symptoms and/or the associated molecular changes. An accurate diagnostic test in early (preclinical) and late stages of the disease is a prerequisite not only for the successful application of future treatments, but also for the correct stratification of individuals for clinical trials.

Polygenic Risk Scores (PRS) are a mathematical aggregate (i.e. a single value) indexing an individual’s relative genetic liability to a trait conferred by hundreds or indeed thousands of risk alleles^3^. The scores are the output of statistical models developed using data from large genome-wide association studies (GWAS). PRS analysis has been widely adopted as a tool for measuring common variant liability in coronary artery disease, schizophrenia, AD, diabetes and cancer^4-9^. Furthermore, there have been efforts to develop the use of PRS as a diagnostic tool (i.e. as a biomarker) for early identification of people at an increased risk manifestation of clinical disease^9^.

In AD PRS have been used to predict lifetime risk of AD development^4, 10, 11^, yielding Area Under the Curve (AUC) estimates in identifying individuals with pathologically confirmed AD vs. controls of ∼82-84%^11^, including the *APOE* locus. In addition, the sensitivity (true positives) increases to ∼90% for PRS extremes^12^. Thus far, efforts to exploit GWAS associations to identify pathological mechanisms underpinning AD have met with varying success^13^, but immune response, lipid metabolism, regulation of Aß formation and cholesterol metabolism, have been identified as likely to be key disrupted biological mechanisms^14^ and macrophages and microglia as likely key drivers of pathology^15^. As for AD PRS, based on many variants in a cumulative fashion, understanding the underlying molecular or biological mechanisms that comprise the polygenic component in AD through gene-expression data, have not been explored before.

To address the paucity of knowledge with respect to the downstream molecular consequences of genetic liability to AD and to understand the biological mechanisms that are likely to be impacted upon by increased liability, we analysed the differential gene-expression from bulk RNA sequencing in the MayoRNAseq publicly available dataset with respect to PRS. We also compare the findings to a case/controls differential gene-expression analysis (Figure 1). This offers a potential to extend the clinical utility of PRS beyond diagnosing individuals at high risk of AD by pointing to putative causal processes at the molecular level.

**Figure 1.**
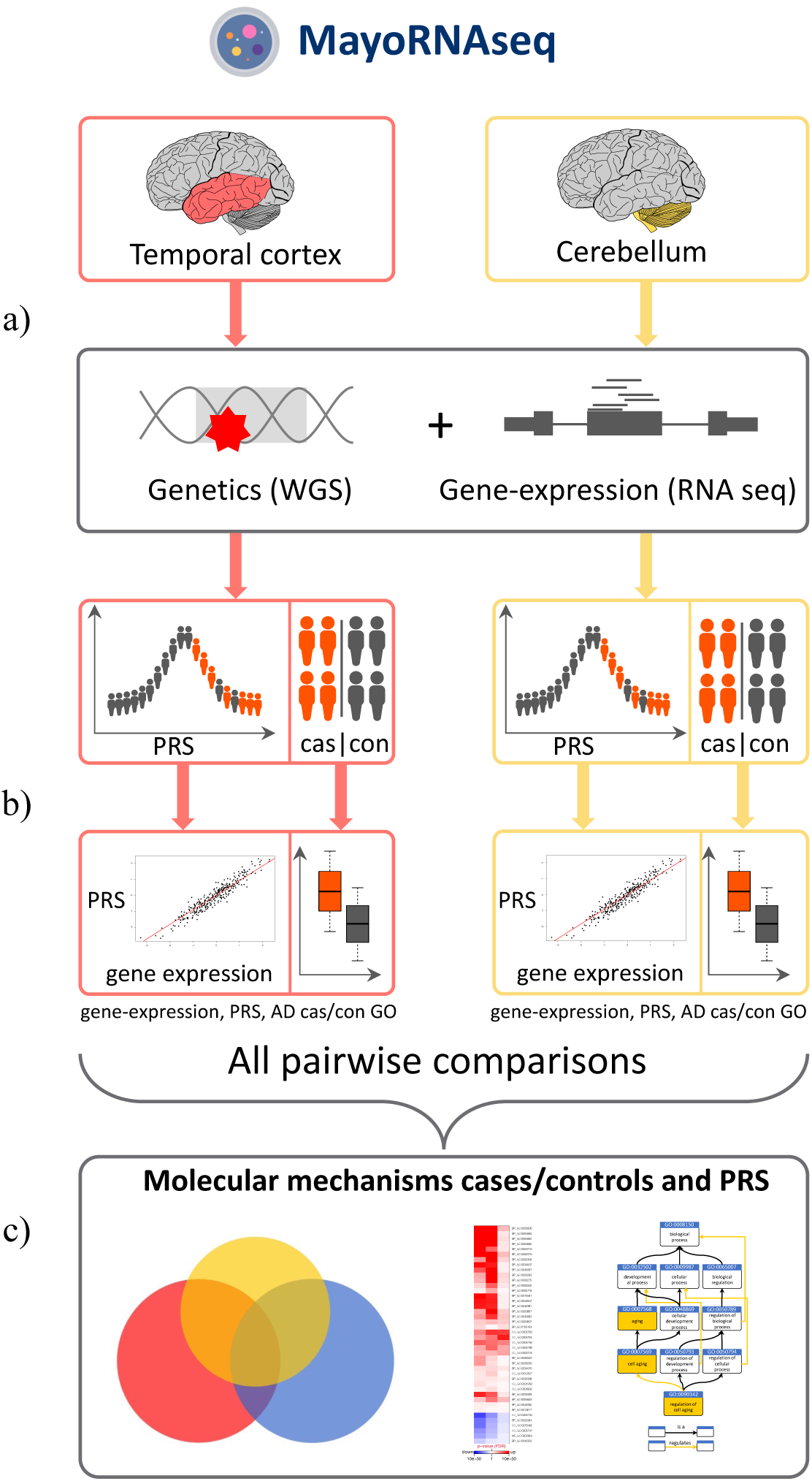
Experimental flowchart. a) MayoRNAseq comprise individuals with matched genetic (WGS) and gene-expression data (bulk brain RNA-seq) from two brain samples: cerebellum and temporal cortex. b) Differentially expressed genes were derived separately for case/controls and PRS. c) Gene-ontology enrichment analysis was performed separately for both case/control and PRS outcomes and compared pairwise across all analysis.

## 2. Materials and Methods

### 2.1. Sample data

We used the MayoRNAseq study (MayoRNAseq)^16^, part of the Accelerated-Medicine Partnership (AMP-AD). MayoRNAseq is a post-mortem brain cohort sample of individuals with a neuropathological diagnosis of AD, progressive supranuclear palsy, or pathological ageing, and elderly controls. The MayoRNAseq human brain-derived samples comprise temporal cortex and cerebellum tissues. The number of samples and other descriptors can be found in Suppl. Table 1.

### 2.2. WGS, RNA-seq and metadata

WGS recalibrated vcf files (*.recalibrated_variants.vcf.gz) were downloaded from the AMP-AD consortium website (https://www.synapse.org/#!Synapse:syn22264775). RNA-seq bam files were also downloaded from the ConsortiumStudies (https://www.synapse.org/#!Synapse:syn9702085). All available metadata for the MayoRNAseq as part of AMP-AD were combined and WGS and RNA-seq individual identifiers were matched according to the manifest files.

### 2.3. Alignment of RNA-seq to human reference genome build 38

The original bam files were converted to fastq (GATK picard-tools 1.60; SamToFastq) and aligned to human reference genome build 38 (GRCh38.98; gtf and fasta http://ftp.ensembl.org/pub/release-98/gtf/homo_sapiens/) using STAR aligner^17^ (v2.7.1a). Duplicated reads were marked using picard-tools 1.60 (MarkDuplicates) and RG groups populated using samtools^18^ (v1.9).

### 2.4. RNA-seq quality control (QC)

RNA-seq QC of the aligned GRCh38.38 bam files was performed using RNA-SeQC 2.3.5^19^. Individual RNA-SeQC measures were converted to percentage and means and standard deviations were calculated for these measures separately within the two tissue MayoRNAseq samples, cerebellum and temporal cortex. Samples were excluded from further analysis if a specific RNA-SeQC measure for an individual RNA-seq sample was 4 standard deviations away from the mean of the distribution (≥ or ≤4 standard deviations considered for different measures). More information on the measures used is provided in the supplementary materials (Suppl. Table 2).

Read counts per gene per sample were derived using htseq-count (v0.11.2). Samples were removed from further analysis if they had 0 read counts across all genes. Genes were removed from further analysis if they had 0 read counts across all samples. Read counts were normalised using the trimmed mean of M-values (TMM) normalization method in the R/bioconductor package edgeR^20^ (v3.34.0) to estimate scaling factors and to adjust for differences in library sizes. Genes were excluded from further analysis if TMM values were <0.5 in 50% of the MayoRNAseq samples.

Raw gene counts derived from htseq-count for the remaining samples were normalized using Conditional Quantile Normalization (CQN)^21^ (R/bioconductor package cqn v1.38.0) to use for principal component analysis (CQN-normalised counts; y+offset). Gene exon lengths and GC content were calculated from the gtf and fasta files using custom built programs.

### 2.5. VCF QC

Individual vcf files were converted and merged with PLINK ^22^ (PLINK v2.00a2.3) and bi-allelic variants were kept. For ethnicity estimates we also downloaded phase3 1000 Genomes Project reference data^23^ from PLINK’s website (https://www.cog-genomics.org/plink/2.0/resources#1kg_phase3) and converted to PLINK format as well as removed duplicates with respect to genomic position. Only variants that were present in the 1000 Genomes phase3 were kept for further analysis (variants matched by chromosome and position). The chromosome and position of the remaining variants were converted with respect to the human genome reference GRCh38 using the UCSC web-based liftover tool (http://genome.ucsc.edu/cgi-bin/hgLiftOver). Variants with Hardy-Weinberg equilibrium (p≤1×10^−06^), missingness (≥0.05) and minor allele frequency (≤0.01) were excluded from further analysis. The remaining filtered variants were combined with the 1000 Genomes phase3. Ancestry was estimated using Principal Component Analysis (PCA) in PLINK2 (--pca --maf) by plotting the first two eigenvectors and samples were excluded from further analysis if a sample deviated from the 1000 Genomes EUR cluster (Suppl. Figure 1c). In order to estimate individual sex, we removed the pseudo-autosomal region of the X chromosome and calculated inbreeding coefficients (F) using PLINK (--check-sex). Individuals with F≤0.2 were deemed females and F≥0.8 males.

Genetic relationship between the samples (pairwise identity by descent (IBD)) was determined using PLINK (--genome full --min 0.1). Samples that had PI-HAT≥0.22 were considered duplicates, or first-degree relatives (including identical twins). All such pairs of samples were excluded from further analysis.

### 2.6. Matching RNA-seq to VCF samples

Matching individual RNA-seq with the VCF samples was done using verifyBamID^24^ (v1.1.3). A matched sample comprising RNA-seq and WGS vcf files from the same individual was done based on the IBD coefficients from verifyBamID; a matched sample was deemed IBD≥0.8. Only matched RNA-seq with WGS vcf files were included for further analysis.

### 2.7. AD diagnosis and APOE status

The MayoRNA-seq dataset included diagnostic status in the RNA-seq covariates file from the AMP-AD knowledge portal. We excluded samples with progressive supranuclear palsy and pathological ageing, retaining only data from samples with a label of AD or control. To assign APOE status we used rs429358 and rs7412. The ambiguous double heterozygotes were coded as E2/E4 (Suppl. Table 3).

### 2.8. RNA-seq differential gene-expression

We used R/bioconductor package *DESeq2* (v.1.32.0)^25^ with raw htseq-counts with age at death, sex and *APOE* status as covariates (*DESeq2* model matrix: design=∼age_at_death+sex+APOE_status+diagnosis for case/control analysis and design=∼age_at_death+sex+APOE_status+PRS for PRS; log fold changes and p-values are returned for the last variable in the design matrix). To account for multiple hypotheses testing the Benjamini-Hochberg false discovery rate was used (FDR).

### 2.9. Gene Ontology

The Wilcoxon rank sum test, as implemented in Catmap^26^, was used to test for significant enrichment of Gene Ontology (GO) categories (differential gene-expression associated with PRS and separately for differential gene-expression with case/controls) using custom built gene-GO gene association (Suppl. Materials and Methods). Ranks of genes were based on the p-value from the significance of the differential gene-expression (from *DESeq2*). For all tests, three lists were derived comprising (1) differentially expressed genes based on p-value only (termed no-direction), (2) the most differentially up-regulated (p-value and log-fold) genes at the top of the list and most differentially down-regulated genes (log-fold< 0) at the bottom of the list (termed up-regulated) and (3) the most differentially down-regulated (log-fold< 0) genes at the top of the list and most differentially up-regulated genes (log-fold> 0) at the bottom of the list (termed down-regulated). Gene lists 2) and 3) are inverted copies of each other. We used random null as the null hypothesis. Even though a random null is not as good approximation as compared to sample label permutations^26^, it was deemed computationally unfeasible to perform sample-label permutations. Nevertheless, we also performed a functional GO enrichment analysis using a separate method (*topGO*^27^; v2.44.0) with the three sets of ranked list of genes using the classic algorithm with the ks statistic (Kolmogorov-Smirnov test) and compared the results with Catmap. To account for multiple hypotheses testing the Benjamini-Hochberg false discovery rate was used. Statistical significance of overlaps of GOs between two experiments (e.g. Catmap vs. topGO, significant GO terms associated with PRS vs. significant GO terms from a case/control analysis) was determined using a hypergeometric test (including Biological Process (BP), Cellular Component (CC) and Molecular Function (MF) GO terms) and profile similarity by using a paired rank-based test for association based on Spearman’s *rho* (gene-expression and GO).

For clustering of statistically significant GO terms we used semantic similarity (*GOSemSim*^28^) with *Rel* information content measure and classical multidimensional scaling (CMD; *cmdscale* package in *R*, k=2) separately for BP and CC GO terms as semantic similarity can only be performed within BP or CC. The most representative (manually curated) GO term was chosen as the name for describing CMD clusters.

### 2.10. PRS calculations

To generate polygenic risk scores for the MayoRNAseq dataset we used the summary statistics from the clinically assessed case/control study on AD^14^, excluding AMP-AD samples that are part of that GWAS^14^. We chose to use Kunkle *et al*.^14^ AD GWAS, as it does not include the UK Biobank summary statistics, where cases are defined via family history (AD proxies^29^) and controls are not screened for AD. PRS were calculated using PLINK for pT≤0.1 (p-value threshold) on LD-clumped SNPs by retaining the SNP with the smallest p-value excluding SNPs with *r*^2^>0.1 in a 1000kb window. We chose to use the simplest clumping and thresholding approach as the simplicity guarantees the transparency (included SNPs and weights are known) and the prediction accuracy of AD by the PRS is similar for the majority of methodologies as previously described^30^. All derived scores were adjusted for 5 consecutive principal components then standardised within the MayoRNAseq samples (mean and standard deviation).

## 3. Results

### 3.1. Sample numbers after QC

After our quality control (genotypes and RNA-seq), there were 288 samples with matched genetic and RNA-seq samples in the MayoRNAseq dataset (170 genetically unique individuals; Suppl. Table 1).

### 3.2. AD case/control differential gene-expression and GO enrichment

Differentially expressed genes were derived using *DESeq2* separately for the two tissue samples in the MayoRNAseq (temporal cortex and cerebellum), including covariates for age at death, sex and *APOE* status. There were >5,000 differentially expressed genes after correction for multiple hypothesis testing in both cerebellum (∼8,000) and temporal cortex (∼5,000; Suppl. Data 1 & 2) with a statistically significant overlap of differentially expressed genes between the two tissues (Suppl. Figure 2). There was no statistically significant enrichment of AD-associated GWAS risk genes in any of the three gene lists (p=0.97 and p=0.31 for cerebellum and temporal cortex respectively for genes based only on p-value (no-direction); p=0.42 and 0.06 for up-regulated (order by p-value and logfc); p=0.58 and p=0.94 for down-regulated; list of AD GWAS risk genes given in Suppl. Data 3 and description in Suppl. Materials and Methods).

We performed GO enrichment analysis (biological process (BP), cellular component (CC) and molecular function (MF)) using the three sets of differential expression gene lists, that is no-direction (based on p-value only), up-regulated and down-regulated (log-fold change and p-value). There was a statistically significant overlap of significantly enriched GO terms (separately for all three gene lists) between the two tissues (Suppl. Figure 3a-e) in addition to a significant GO rank profile similarity (Suppl. Figure 3f-h). This suggests that both tissues share an overall statistically significant similarity in terms of disrupted biological pathways with respect to a case/control analysis. It is of note that overlap of GO and testing for profile similarity achieved much stronger statistical significance in the up and down-regulated significant GO terms as compared to the no-direction results (gene order based on p-value only).

The statistically significant GO terms from both tissues (**no-direction** gene-list; p-value only) were combined and clusters were derived using semantic similarity (BP and CC). This was done to reduce the complexity and functional redundancy of GO terms. Significantly disrupted biological processes included response to stimulus, regulation of signal transduction, cell motility and metabolism, aerobic respiration, differentiation, organelles (Golgi Apparatus, endoplasmic reticulum (ER), mitochondria), oxidoreductase complex, cell cycle, regulation of cell death (Suppl. Figure 4).

We also performed the same semantic similarity clustering separately for the up-regulated and down-regulated GO terms. Significantly disrupted **up-regulated** biological processes included regulation of metabolism (including lipid and cholesterol), stress and immune response, signalling, DNA repair, differentiation/morphogenesis/development, organelles (Golgi Apparatus, ER, mitochondria), senescence, neuronal cells (Suppl. Figure 5). Significantly disrupted **down-regulated** biological processes mainly included cellular respiration such as mitochondrial electron transport, respiratory chain complexes, mitochondrial membrane, etc. (Suppl. Figure 6). GO-terms in all semantic similarity clusters (both up and down-regulated analysis) were from both tissue samples, replicating the statistically significant profile GO similarity and overlap of GO terms, suggesting limited overall brain region specificity.

GO enrichment analysis showed that several biological pathways previously implicated from GWAS^14^ in AD were significantly enriched in the AD case/control differential gene-expression analysis (Suppl. Data 4-5, Figure 2a and Figure 5a), although we have not formally tested if this is statistically significant, thus it could be a chance finding. Significantly **up-regulated** GO terms included **immune system processes** (GO:0002376, p=1.13e-06 and p=4.09e-43 for cerebellum and temporal cortex respectively), **response to lipids** (GO:0033993, p=7.41e-03 and p=1.05e-27), **inflammatory response** (GO:0006954, p=4.87e-02 and p=5.83e-15), **endosome** (GO:0005768, p=6.27e-07 and p=2.30e-11), **regulation of cell death** (GO:0010941, p=1.78e-08 and p=1.26e-32), **regulation of neuron death** (GO:1901214, p=6.46e-03 and p=1.32e-04). There was also evidence for the involvement of glial cells in the temporal cortex (up-regulated GOs, **glial cell projection** GO:0097386 p=1.62e-04, **astrocyte projection** GO:0097449 p=6.55e-04, **regulation of microglial cell activation** GO:1903978 p=1.24e-02), but not in cerebellum. In addition, we were only able to confirm such previous AD GWAS-derived disrupted biological pathways by sorting (log-fold change and p-value) or in other words using the direction of effect of the genes (mostly up-regulated and mostly down-regulated at the top of the gene lists), but not based on p-value only (no-direction). For example, immune system process (GO:0002376) was not significantly enriched GO-term in the no-direction (genes sorted by p-value only; FDR p=1 and p=8.07e-01 in cerebellum and temporal cortex respectively), but it was significantly enriched in both cerebellum and temporal cortex in the up-regulated GO-terms analysis.

**Figure 2.**
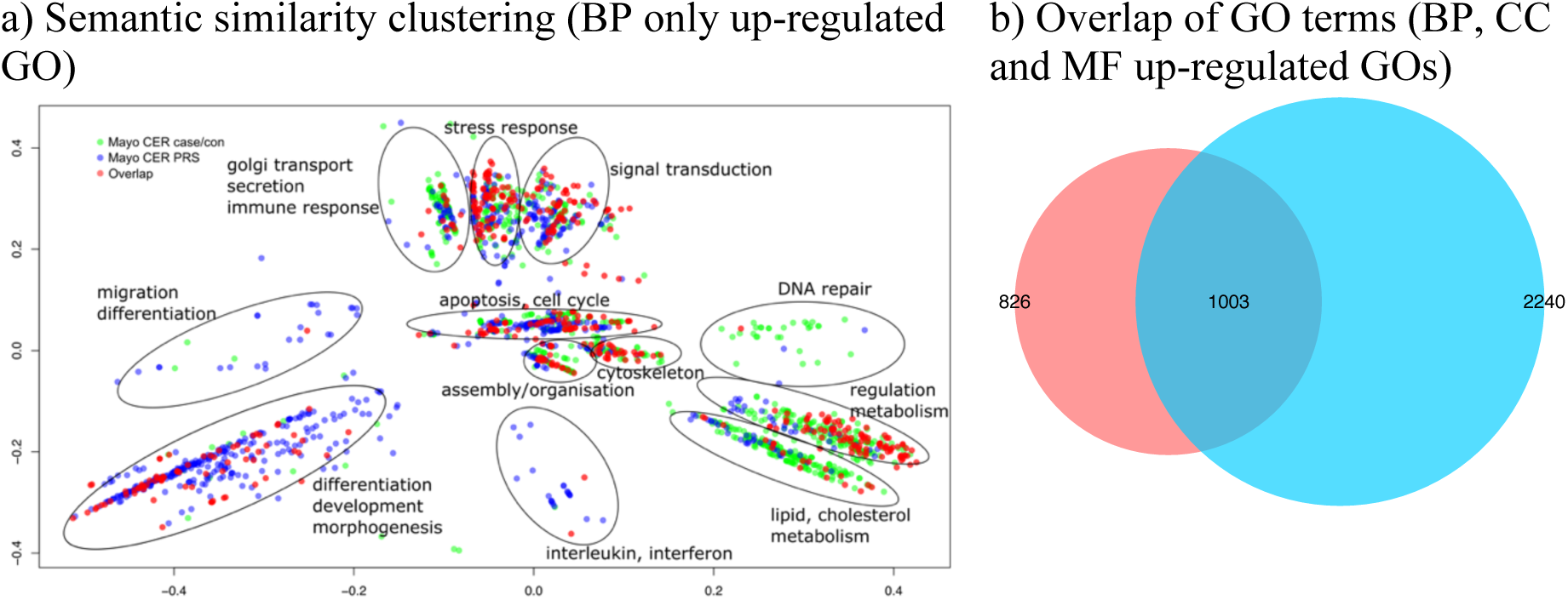
Semantic similarity clustering of up-regulated statistically significant GO terms in both cerebellum and temporal cortex (case/control analysis) a) X and Y axes represent classical multidimensional scaling (CMD) dimension 1 and 2. All GO terms p≤0.05 FDR. Green dots represent significant GO terms from the case/control analysis of cerebellum, Blue dots represent significant GO terms from the case/control analysis of temporal cortex, Red dots represent significant GO terms overlapping in case/control analysis of cerebellum and temporal cortex. Cluster labels were manually curated based on the most common GO term in the cluster. b) Proportional Venn diagram. Numbers represent significant GO terms (FDR) in the two lists with the middle number representing the number of genes that overlap. Red colour represents cerebellum, blue-temporal cortex. hypergeometric test p=4.11e-287.

We also performed the GO-terms enrichment analysis using a separate enrichment method (*topGO*^27^) with the same three gene lists (no-direction, mostly up-regulated and mostly down-regulated at the top). The results from both Catmap and topGO (paired GO ranks) display extremely similar rank profile of GO-terms with r^2^ ranging from 0.65-0.8 (Suppl. Figure 7).

### 3.3. PRS differential gene-expression and GO enrichment

Similarly, to the case/control differential gene-expression analysis, for each gene we derived differentially expressed genes associated with PRS using *DESeq2* separately for the two tissue samples in the MayoRNAseq, including a covariate for age at death, sex and *APOE* status.

There were three and 351 genes differentially expressed genes in the cerebellum and the temporal cortex respectively following an FDR correction for multiple hypothesis testing (Suppl. Data 6). There were fewer differentially expressed genes in cerebellum as compared to temporal cortex associated with PRS, in contrast to the fewer differentially expressed genes in the temporal cortex as compared to cerebellum in the case/control analysis. Due to few genes being differentially expressed in cerebellum, we performed an overlap of the top 300 genes in both tissue samples. There was a statistically significant overlap of genes in both datasets in the same direction (including a significant rank correlation of all genes; Suppl. Figure 8).

There was also a statistically significant enrichment of previous AD-associated GWAS risk genes (Wilcox rank-sum test p=2.99e-02; not corrected for multiple hypothesis testing) in the temporal cortex no-direction gene list (ordered by p-value only), but not in cerebellum (p=2.44e-01). The top ranked 15 genes in the temporal cortex also found in AD GWAS hits were *HAVCR2, MS4A6A, INPP5D, ECHDC3, SPI1, ADAMTS4, ADAMTS1, CR1, IL34, PICALM, HLA-DRB1, CD33, APH1B, FERMT2*, and *PLCG2*, although only *HAVCR2* and *MS4A6A* (p=1.61e-02, beta=0.21 and p=3.72e-02; beta=0.29), passed FDR correction. In addition, there was a statistically significant enrichment of AD-associated GWAS genes in the up-regulated gene list in temporal cortex (p=1.22e-05 and p=0.49 for temporal cortex and cerebellum respectively), but not in the down-regulated gene list for both temporal cortex and cerebellum (p=0.5 and p=0.99). This suggests that overall GWAS-hits are on average ranked significantly higher in the temporal cortex gene expression list in the PRS analysis than expected by chance alone and these are more likely to be up-regulated than down-regulated (only *IL34* was down-regulated among the top 15 GWAS-hits). Furthermore, in temporal cortex, higher AD PRS was associated with increased gene-expression of 52 out of 75 AD GWAS associated genes (Fisher’s exact test p=2.79e-04; 10319 up and 11071 down-regulated among all genes).

There was a statistically significant overlap of significantly enriched GO terms (separately for all three gene lists) between the two tissues (Suppl. Figure 9a-e) in addition to a significant GO rank profile similarity (Suppl. Fig9f-h). This suggests that both tissues share an overall similarity in terms of disrupted biological pathways with respect to PRS.

The statistically significant GO terms from both tissues (no-direction gene-list; p-value only) were combined and clusters were derived using semantic similarity. Significantly disrupted biological processes (**no-direction** gene list) included immune response, stress response, regulation of metabolism, transport and signalling, aerobic respiration, organelles (Golgi apparatus, ER, mitochondria), oxidoreductase complex, cell cycle, regulation of cell death (Suppl. Figure 10). Nevertheless, GOs in immune-related clusters (i.e. immune response, regulation of T/B cells and interferon/interleukin) were statistically significant only in temporal cortex (Suppl. Figure 10a), but not in cerebellum.

The semantic similarity clustering was also performed separately for the up-regulated and down-regulated GO terms. Significantly disrupted **up-regulated** biological processes included regulation of metabolism (including fatty acids and cholesterol), stress and immune response (adaptive and innate), signalling, DNA repair, differentiation/morphogenesis/development, organelles (Golgi Apparatus, ER, mitochondria), senescence and neuronal cell death, neuronal cells (Suppl. Figure 11 and Figure 3a). Significantly disrupted **down-regulated** biological processes mainly included cellular respiration such as mitochondrial electron transport, respiratory chain complexes, mitochondrial membrane, mitochondrial ATP synthesis, metabolism, neuronal processes such as neurotransmitter secretion/transport, neuron projection, postsynaptic membrane (Suppl. Figure 12). The semantic similarity clusters comprised up-regulated GO terms from both tissues (semantic similarity Suppl. Figure 11), but there were notable differences in the down-regulated GOs, suggesting tissue specificity. All synaptic-associated GO-terms were found to be significantly down-regulated in temporal cortex, but up-regulated cerebellum. These include synaptic/neuronal processes such as synaptic signalling, synaptic and pre/postsynaptic membranes, regulation of synaptic plasticity, synaptic vesicle, neurotransmitter secretion, glutamatergic synapse, etc. (Figure 3c and Figure 5a).

**Figure 3.**
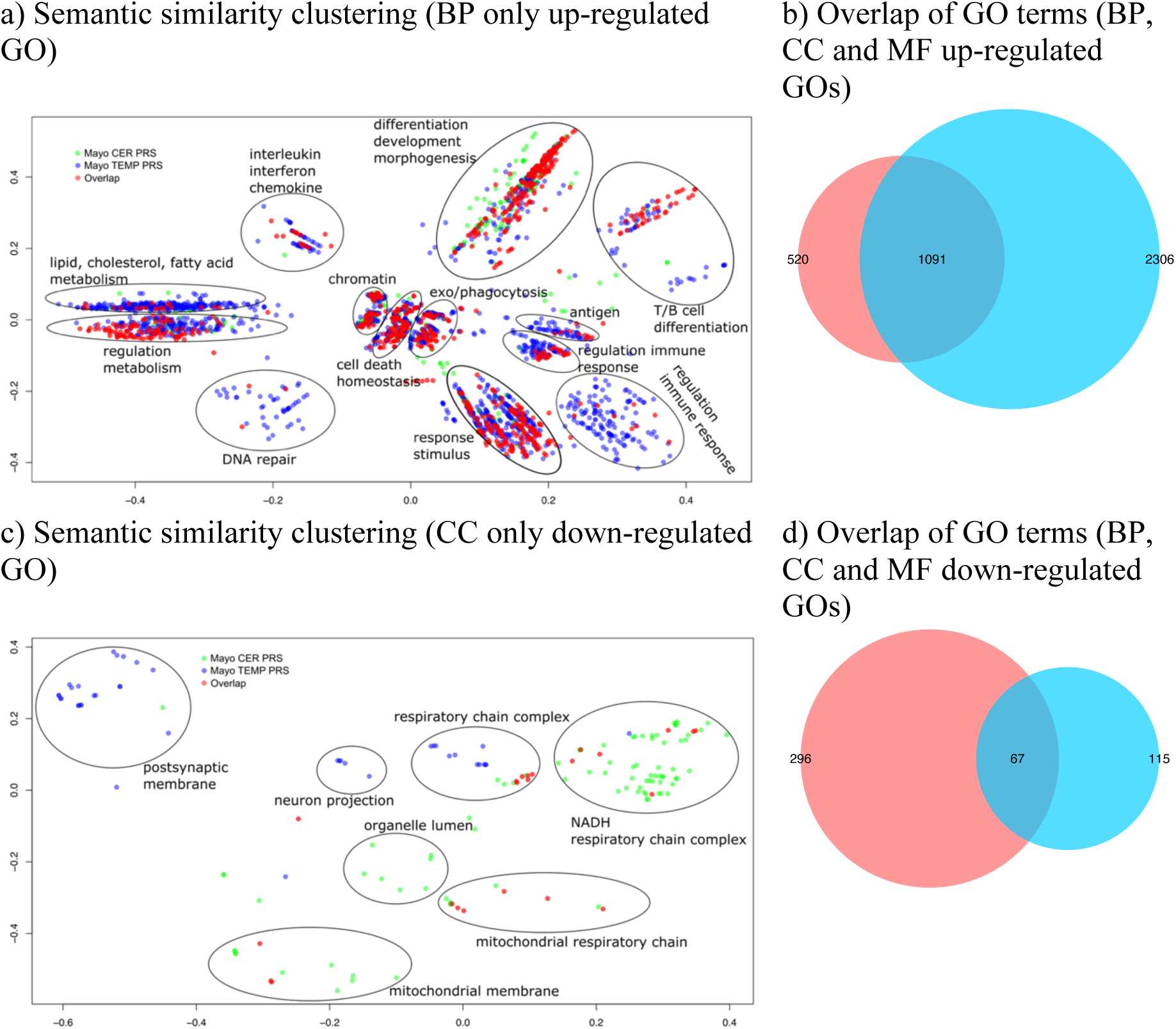
Semantic similarity clustering of up-regulated statistically significant GO terms in both cerebellum and temporal cortex (PRS analysis) a) and c) X and Y axes represent classical multidimensional scaling (CMD) dimension 1 and 2. All GO terms p≤0.05 FDR. Green dots represent significant GO terms from the PRS analysis of cerebellum, Blue dots represent significant GO terms from the PRS analysis of temporal cortex, Red dots represent significant GO terms overlapping in PRS analysis of cerebellum and temporal cortex. Cluster labels were manually curated based on the most common GO term in the cluster. b) Proportional Venn diagram. Numbers represent the significant GO terms (FDR) in the two lists with the middle number representing the number of genes that overlap. Red colour represents cerebellum the blue temporal cortex. b) hypergeometric test p<1e-300; c) hypergeometric test p=6.85e-65.

Similarly, to the case/control analysis GO enrichment analysis showed that a wide range of previously implicated (from GWAS) biological pathways in AD were also found to be significantly enriched (Suppl. Data 10-12 and Figure 3a and b and Figure 5a), including **immune system processes** (GO:0002376, p= 2.72E-05 and p= 2.67E-98 for cerebellum and temporal cortex respectively), **response to lipids** (GO:0033993, p= 1.44E-07 and p= 1.17E-21), **inflammatory response** (GO:0006954, p= 7.98E-04 and p= 3.24E-29), **endosome** (GO:0005768, p= 2.44E-03 and p= 4.48E-19), **regulation of cell death** (GO:0010941, p= 2.26E-07 and p= 1.12E-27), **regulation of neuron death** (GO:1901214, p= 2.55E-03 and p= 2.96E-02). There was also some evidence for the involvement of glial cells in both tissues (up-regulated GOs, **glial cell projection** GO:0097386 p=1.95e-03 and p=3.48e-02 for cerebellum and temporal cortex respectively, **astrocyte activation** GO:0048143 p=3.66e-02 and p=1.26e-02, **microglial cell activation** GO:0001774 p=4.09e-02 and p=3.09e-07).

Similarly, to the case/control GO analysis, the results from both Catmap and topGO (paired GO ranks) displayed extremely similar rank profile of GO-terms with r^2^ ranging from 0.65-0.81 (Suppl. Figure 13).

### 3.4. Molecular mechanisms shared/different between cases/controls and PRS with respect to differential gene expression

We compared the differential expression results in terms of genes from the case/control and PRS analyses for cerebellum and temporal cortex respectively. There was no statistically significant overlap of differentially expressed genes in cerebellum (Suppl. Figs. 14 & 15), but there was a statistically significant overlap of differentially up and down-regulated genes in the temporal cortex (Suppl. Figure 16).

Contrary to the results with respect to overlap of differentially expressed genes, the overlap of GO terms for both cerebellum and temporal cortex showed remarkable similarity in terms of both overlap of significantly disrupted GOs and rank profiles in all three gene lists (no-direction, most up-regulated at the top and most-downregulated and top; Suppl. Figs. 17 and 18), although fewer GOs overlapped if no-direction of gene effect was used.

The statistically significant GO terms from both tissues (**no-direction** gene-list; p-value only) from the case/control and PRS analyses were combined and clusters were derived using semantic similarity, separately for cerebellum and temporal cortex. There were fewer significantly disrupted GO terms in the case/control analysis as compared to PRS (57 vs. 389 in cerebellum and 264 and 695 for temporal cortex for the case/control and PRS respectively; Suppl. Data 4a, 5a, 7a and 8a). The only processes that were in common in cerebellum were GOs related to organelles and metabolic processes (Suppl. Figure 19). Similarly, the commonly disrupted biological processes in temporal cortex were extracellular space/structure, organelles, response to stimulus/lipids, signal transduction (Suppl. Figure 22). Most of the semantically similar clusters of **up/down-regulated** GOs in cerebellum with respect to case/controls and PRS comprised GOs from both analyses (case/controls and in response to PRS), suggesting similarly disrupted biological processes with very few differences (Suppl. Figs. 20 & 21). Differences included significantly down-regulated biological processes found only in response to PRS such as, WNT/NF-kappaB signalling, rRNA processing, protein import in mitochondria (Suppl. Figure 21) and significantly up-regulated processes only found in case/control analysis such as, histone acetyltransferase complexes (Suppl. Figure 20).

Similarly to cerebellum, most of the up-regulated semantically similar clusters in temporal cortex (case/control vs. PRS) have GO terms from both case/control and PRS analysis, suggesting little differences in terms of significantly disrupted up-regulated biological processes (Suppl. Figs. 23 and Figure 4a). This was in contrast to down-regulated terms that showed differences. These included mainly neuronal/synaptic down-regulated processes only found in response to PRS as compared to case/control analysis such as, neuronal plasticity, synaptic signalling/transmission, neurotransmitter levels and secretion, post/pre-synaptic membrane, glutamatergic and GABA-ergic synapse (Suppl. Figure 24 and Figure 4b).

**Figure 4.**
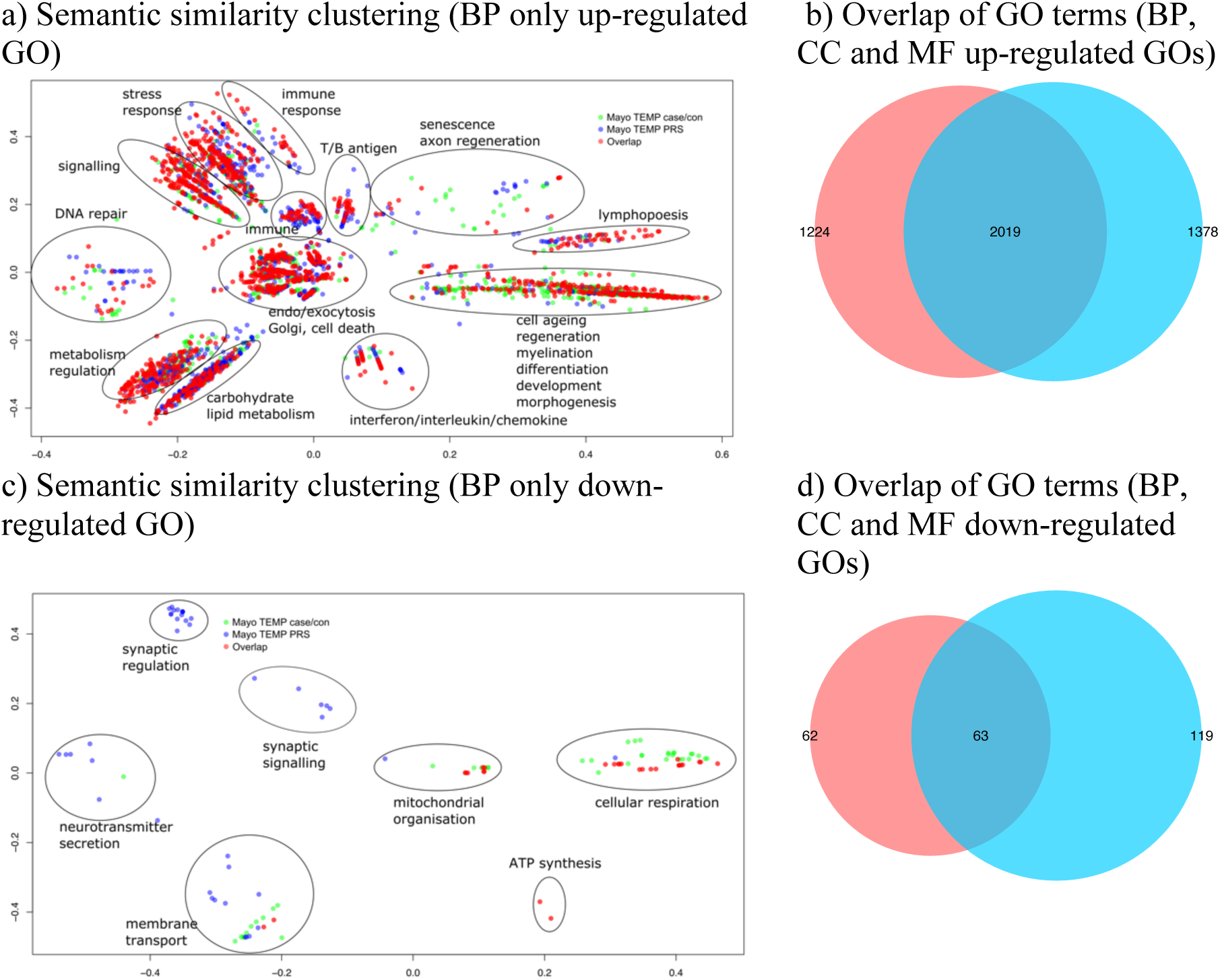
Semantic similarity clustering of up and down-regulated statistically significant GO terms in temporal cortex (case/control vs. PRS analysis) a) and c) X and Y axes represent classical multidimensional scaling (CMD) dimension 1 and 2. All GO terms p≤0.05 FDR. Green dots represent significant GO terms from the case/control & PRS analysis of temporal cortex, Blue dots represent significant GO terms from the case/control & PRS analysis of temporal cortex, Red dots represent significant GO terms overlapping in case/control analysis of cerebellum and temporal cortex. Cluster labels were manually curated based on the most common GO term in the cluster. b) and d) Proportional Venn diagram. Numbers represent the significant GO terms (FDR) in the two lists with the middle number representing the number of genes that overlap. Red colour represents case/control the blue PRS analyses. hypergeometric test for b) p<1e-300; hypergeometric test for d) p=3.04e-93.

We parsed all the GO-terms from all the analysis (case/control and PRS in cerebellum and temporal cortex) using search terms from previously reported molecular mechanisms disrupted in AD^14, 31^. The search terms were grouped in eight categories, aging/senescence, death/apoptosis, neuron/synapse, glial cell populations, amyloid, immune response, stress response, lipid/cholesterol/fatty acid metabolism. GO-terms matching any of the search terms and are statistically significant in at least one analysis were retained and sorted by the mean - log10 p FDR across all the analyses. The most statistically significant categories were immune and stress response, asserting an important role of the immune system in the development of AD^14^ (Figure 5a). The least significant were glial cell populations and amyloid. This analysis does not take into account the overlap of genes within different Gos and the overall redundancy of GO terms. It is of note that in all the differential gene-expression analyses (case/control and PRS) we included age of death as a fixed covariate and despite this, ageing GO term is still a significant molecular mechanism associated with the development of AD.

**Figure 5.**
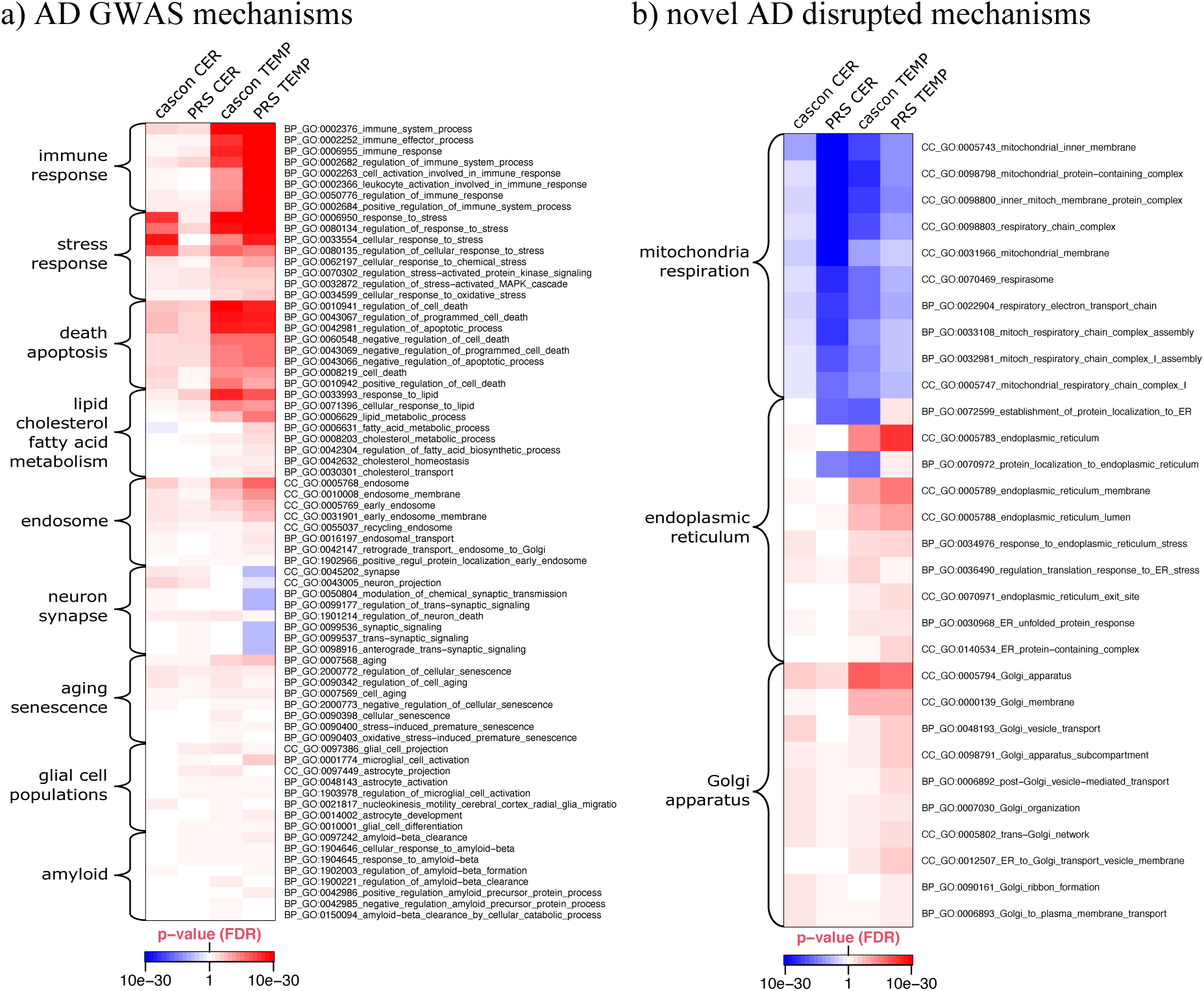
AD GWAS and novel mechanisms statistically significant in MayoRNAseq temporal cortex and cerebellum (case/control & PRS) Heatmap of GO terms that are statistically significant in at least one dataset (cas/con & PRS MayoRNAseq temporal cortex and cerebellum). cascon Case/control analysis; CER cerebellum; TEMP temporal cortex. Heatmap p-values are capped at 10e-30. blue colour represents down-regulated GOs and red-colours represent up-regulated GOs. All full GO term names from the up and down-regulated GO term results were searched using stress, immun, neuro/synap, death/apoptosis, lipid/cholesterol/fatty, aging/senescence, glia/astrocyte, abeta, endosome, golgi, reticulum and mitochond/respir and GO terms selected if FDR p-value was <0.05. GO terms within each category were ordered by mean -log10 p and the top 8 selected for visualisation (3 for lipid and cholesterol and 2 for fatty acid metabolism)

Even though, the most significantly disrupted AD GWAS-associated molecular mechanisms were immune/stress response and death/apoptosis, there were other statistically significant GO-terms that have not been reported associated with AD previously and were shared between the case/control and PRS analyses in cerebellum and temporal cortex. These included variety of respiration-related processes (e.g. respiratory electron transport chain, mitochondrial inner membrane), Golgi apparatus and endoplasmic reticulum (Figure 5b).

## 4. Discussion

We performed an integrative analysis of the transcriptomics using case/control and genetic liability paradigms. The main aim of the study was to try to understand the biological correlates of elevated common variant liability to AD, and their relationship with these associated with AD *per se*.

Our overall findings suggest that disrupted biological pathways associated with affected status and increased PRS show remarkable profile similarities with respect to biological pathways derived from gene-expression (bulk brain-derived RNA-seq). In temporal cortex, we found evidence for a modest degree of similarity with respect to genes that are differentially expressed in AD cases compared to controls, and those are associated with increased PRS results. However, the degree of similarity between case status and elevated PRS was much stronger at the level of the GO-term enrichments for differentially expressed genes. This suggests a disease heterogeneity in terms of changes in the gene-expression of individual genes^32^, but nevertheless a convergence in terms of disrupted disease biological mechanisms underlying AD. Crucially, this also suggests that both a case/control and PRS classifications elucidate similar molecular mechanisms. There was some evidence for tissue specificity for the associations with PRS, higher PRS being associated with down-regulation of neuronal process genes in temporal cortex, but up-regulation of the same categories in cerebellum. In contrast, there was limited tissue specificity when the dataset was analysed as a case/control sample.

Our gene ontology analysis of differential gene-expression in cases versus controls shows a degree of convergence with analogous analyses of GWAS studies, ^14, 33, 34^, highlighting **immune** (both adaptive and innate) and **stress response, lipids, fatty acids** and **cholesterol metabolism, endosome** and **cellular/neuronal death**. Our results also suggest a significant involvement of previously less well characterised processes in AD. These include the involvement of **cellular structures** (ER, ER stress, Golgi, actin cytoskeleton, lamellipodium) and **cellular mitochondrial respiration** and **secretion** (exocytosis and endocytosis). Most of the AD GWAS implicated loci are non-coding^14, 35^ and choosing the closest gene to an index variant could miss genes that are further away or miss other regulatory mechanisms. Therefore we did not expect to find enrichment of GWAS hits (closest genes) among the differentially expressed genes, although some SNPs have been shown to be directly related to AD^36^. Nevertheless, there was a significant enrichment of differentially expressed genes in the temporal cortex associated with PRS. Thus, some of the putative GWAS implicated genes, as defined as those closest to the associated index SNP at the locus are also likely to show a differential gene-expression in relationship with PRS in temporal cortex. Tissue specificity is also likely to account for some of the differences. The top ranked genes among the differential expression gene list include *HAVCR2, MS4A6A, INPP5D, ECHDC3, SPI1, ADAMTS4, ADAMTS1, CR1, IL34, PICALM, HLA-DRB1, CD33, APH1B, FERMT2*, and *PLCG2*, although only *HAVCR2* and *MS4A6A* passed FDR correction. Strikingly, 69% (52/75; p=2.79e-04) of all GWAS implicated genes were up-regulated in response to different PRS among the temporal cortex samples. While it is beyond the scope of this work, this result suggests a potential common regulatory mechanism or mechanisms. *MS4A6A, INPP5D* and *SPI1* have been previously shown to be dysregulated specifically in microglial cells^31, 37^. Furthermore, the GO term microglial cell activation involved in immune response (GO:0002282) was significantly disrupted in temporal cortex with respect to PRS and it comprises *TYROBP, TREM2, GRN* and *IL33. TYROBP* was significantly up-regulated in response to higher PRS in temporal cortex and has been shown as a strategic and causal regulator in several microglial activation signalling cascades and the complement pathway in late onset AD^38^. Even though the gene-expression data we used are brain-derived (cerebellum and temporal cortex) bulk RNA-seq, we found several disrupted GO terms specifically related to glial cells (Figure 5a). Glial and microglia-related GO terms were not the top ranked GO terms, but nevertheless it is remarkable that this signal is detectable in bulk brain-derived RNA-seq.

The strongest GO-terms enriched in all datasets (both case/control and PRS) were the **ER, Golgi apparatus, mitochondria** and associated **mitochondrial respiratory chain complexes**. These cellular structures have received relatively little attention in AD. Nevertheless, both ER and mitochondrial function have been shown to be altered in AD^39-42^. The ER-mitochondria interaction is tightly linked to changes in lipid and cholesterol metabolism pathways^42^, both of which have been found to be significantly disrupted mechanisms in all datasets used in this work. Furthermore, Aß interacts with ER, Golgi apparatus and mitochondria to disrupt their normal function^43^.

Although age is one of the main risk factors for the development of AD, there is little understanding of the molecular mechanisms involved in this relationship. Most of the AD genetic and genomic statistical analysis use age at death or age of onset to account for the differences in chronological age of research participants and ageing is interchangeably used with age. In this study, despite adjusting our differential gene-expression analysis for age at death, we still found the GO term ageing to be enriched for genes that are up-regulated in a case/control and in response to higher PRS. This suggests that on average the gene-expression of ageing-related genes is markedly changed in individuals with AD as compared to controls and with respect to PRS. This indicates that the use of chronological age in the statistical modelling of genetic/genomic data in AD-research could be flawed. Following Horvath’s seminal paper on estimating biological age using an epigenetic clock^44^, AD individuals have indeed been shown to exhibit an accelerated epigenetic clock and the rate might be also different in different brain regions^45^. Thus, constructing such epigenetic clocks in AD individuals could help delineate the difference between ageing and chronological age and provide further understanding of AD development.

Our study is an integrative computational approach of publicly available data from two sources to try to highlight the biological processes associated with PRS in comparison to case/control classification in AD. Our results point to a considerable heterogeneity in terms of changes in gene-expression with respect to case/control design and genes associated with PRS, but a convergence in terms of disrupted biological pathways, including novel and previous GWAS implicated biological process and cellular structures.

## Data Availability

All data produced in the present work are contained in the manuscript

http://dx.doi.org/10.7303/syn2580853

## 5. Funding/Acknowledgements

The project is funded by Sêr Cymru “Rising Star” fellowship awarded to Dr Dobril Ivanov by the European Regional Development Fund (ERDF) through the Welsh Government (CU-147; Dr Dobril Ivanov and Dr Benoit Lan-Leung). Karen Crawford was supported by MRC Centre for Neuropsychiatric Genetics and Genomics (MR/L010305/1). Dr Ganna Leonenko and Emily Baker were supported by the Dementia Research Institute [UKDRI supported by the Medical Research Council (UKDRI-3003), Alzheimer’s Research UK, and Alzheimer’s Society.

We would like to express our gratitude to the Advanced Research Computing at Cardiff University (ARCCA) for the computational support. We thank Joint Programming for Neurodegeneration (MRC: MR/T04604X/1), Dementia Platforms UK (MRC: MR/L023784/2).

## References

1. 2020 Alzheimer’s disease facts and figures. Alzheimer’s & Dementia. 2020;16:391–460.

2. Gordon BA, Blazey TM, Su Y, Hari-Raj A, Dincer A, Flores S, et al. Spatial patterns of neuroimaging biomarker change in individuals from families with autosomal dominant Alzheimer’s disease: a longitudinal study. Lancet Neurol. 2018;17:241–50.

3. Sugrue LP, Desikan RS. What Are Polygenic Scores and Why Are They Important? JAMA. 2019;321:1820–1.

4. Escott-Price V, Sims R, Bannister C, Harold D, Vronskaya M, Majounie E, et al. Common polygenic variation enhances risk prediction for Alzheimer’s disease. Brain. 2015;138:3673–84.

5. International Schizophrenia C, Purcell SM, Wray NR, Stone JL, Visscher PM, O’Donovan MC, et al. Common polygenic variation contributes to risk of schizophrenia and bipolar disorder. Nature. 2009;460:748–52.

6. Khera AV, Chaffin M, Aragam KG, Haas ME, Roselli C, Choi SH, et al. Genome-wide polygenic scores for common diseases identify individuals with risk equivalent to monogenic mutations. Nat Genet. 2018;50:1219–24.

7. Khera AV, Kathiresan S. Genetics of coronary artery disease: discovery, biology and clinical translation. Nat Rev Genet. 2017;18:331–44.

8. Mavaddat N, Michailidou K, Dennis J, Lush M, Fachal L, Lee A, et al. Polygenic Risk Scores for Prediction of Breast Cancer and Breast Cancer Subtypes. Am J Hum Genet. 2019;104:21–34.

9. Udler MS, McCarthy MI, Florez JC, Mahajan A. Genetic Risk Scores for Diabetes Diagnosis and Precision Medicine. Endocr Rev. 2019;40:1500–20.

10. Chaudhury S, Brookes KJ, Patel T, Fallows A, Guetta-Baranes T, Turton JC, et al. Alzheimer’s disease polygenic risk score as a predictor of conversion from mild-cognitive impairment. Transl Psychiatry. 2019;9:154.

11. Leonenko G, Shoai M, Bellou E, Sims R, Williams J, Hardy J, et al. Genetic risk for alzheimer disease is distinct from genetic risk for amyloid deposition. Ann Neurol. 2019;86:427–35.

12. Sims R, Hill M, Williams J. The multiplex model of the genetics of Alzheimer’s disease. Nat Neurosci. 2020;23:311–22.

13. Flint J, Ideker T. The great hairball gambit. PLoS Genet. 2019;15:e1008519.

14. Kunkle BW, Grenier-Boley B, Sims R, Bis JC, Damotte V, Naj AC, et al. Genetic meta-analysis of diagnosed Alzheimer’s disease identifies new risk loci and implicates Abeta, tau, immunity and lipid processing. Nat Genet. 2019;51:414–30.

15. Novikova G, Kapoor M, Tcw J, Abud EM, Efthymiou AG, Chen SX, et al. Integration of Alzheimer’s disease genetics and myeloid genomics identifies disease risk regulatory elements and genes. Nat Commun. 2021;12:1610.

16. Allen M, Carrasquillo MM, Funk C, Heavner BD, Zou F, Younkin CS, et al. Human whole genome genotype and transcriptome data for Alzheimer’s and other neurodegenerative diseases. Sci Data. 2016;3:160089.

17. Dobin A, Davis CA, Schlesinger F, Drenkow J, Zaleski C, Jha S, et al. STAR: ultrafast universal RNA-seq aligner. Bioinformatics. 2013;29:15–21.

18. Danecek P, Bonfield JK, Liddle J, Marshall J, Ohan V, Pollard MO, et al. Twelve years of SAMtools and BCFtools. Gigascience. 2021;10.

19. Graubert A, Aguet F, Ravi A, Ardlie KG, Getz G. RNA-SeQC 2: Efficient RNA-seq quality control and quantification for large cohorts. Bioinformatics. 2021.

20. McCarthy DJ, Chen Y, Smyth GK. Differential expression analysis of multifactor RNA-Seq experiments with respect to biological variation. Nucleic Acids Res. 2012;40:4288–97.

21. Hansen KD, Irizarry RA, Wu Z. Removing technical variability in RNA-seq data using conditional quantile normalization. Biostatistics. 2012;13:204–16.

22. Chang CC, Chow CC, Tellier LC, Vattikuti S, Purcell SM, Lee JJ. Second-generation PLINK: rising to the challenge of larger and richer datasets. Gigascience. 2015;4:7.

23. Genomes Project C, Auton A, Brooks LD, Durbin RM, Garrison EP, Kang HM, et al. A global reference for human genetic variation. Nature. 2015;526:68–74.

24. Jun G, Flickinger M, Hetrick KN, Romm JM, Doheny KF, Abecasis GR, et al. Detecting and estimating contamination of human DNA samples in sequencing and array-based genotype data. Am J Hum Genet. 2012;91:839–48.

25. Love MI, Huber W, Anders S. Moderated estimation of fold change and dispersion for RNA-seq data with DESeq2. Genome Biol. 2014;15:550.

26. Breslin T, Eden P, Krogh M. Comparing functional annotation analyses with Catmap. BMC Bioinformatics. 2004;5:193.

27. Alex A, Rahnenfuhrer J. topGO: Enrichment Analysis for Gene Ontology. R package version 2.44.0. doi:10.18129/B9.bioc.topGO. 2021.

28. Yu G. Gene Ontology Semantic Similarity Analysis Using GOSemSim. Methods Mol Biol. 2020;2117:207–15.

29. Marioni RE, Harris SE, Zhang Q, McRae AF, Hagenaars SP, Hill WD, et al. GWAS on family history of Alzheimer’s disease. Transl Psychiatry. 2018;8:99.

30. Leonenko G, Baker E, Stevenson-Hoare J, Sierksma A, Fiers M, Williams J, et al. Identifying individuals with high risk of Alzheimer’s disease using polygenic risk scores. Nat Commun. 2021;12:4506.

31. Morabito S, Miyoshi E, Michael N, Swarup V. Integrative genomics approach identifies conserved transcriptomic networks in Alzheimer’s disease. Hum Mol Genet. 2020;29:2899–919.

32. Crist AM, Hinkle KM, Wang X, Moloney CM, Matchett BJ, Labuzan SA, et al. Transcriptomic analysis to identify genes associated with selective hippocampal vulnerability in Alzheimer’s disease. Nat Commun. 2021;12:2311.

33. Small SA, Simoes-Spassov S, Mayeux R, Petsko GA. Endosomal Traffic Jams Represent a Pathogenic Hub and Therapeutic Target in Alzheimer’s Disease. Trends Neurosci. 2017;40:592–602.

34. Jones L, Holmans PA, Hamshere ML, Harold D, Moskvina V, Ivanov D, et al. Genetic evidence implicates the immune system and cholesterol metabolism in the aetiology of Alzheimer’s disease. PLoS One. 2010;5:e13950.

35. Jansen IE, Savage JE, Watanabe K, Bryois J, Williams DM, Steinberg S, et al. Genome-wide meta-analysis identifies new loci and functional pathways influencing Alzheimer’s disease risk. Nat Genet. 2019;51:404–13.

36. Zhao T, Hu Y, Zang T, Wang Y. Integrate GWAS, eQTL, and mQTL Data to Identify Alzheimer’s Disease-Related Genes. Front Genet. 2019;10:1021.

37. Tsai AP, Lin PB, Dong C, Moutinho M, Casali BT, Liu Y, et al. INPP5D expression is associated with risk for Alzheimer’s disease and induced by plaque-associated microglia. Neurobiol Dis. 2021;153:105303.

38. Zhang B, Gaiteri C, Bodea LG, Wang Z, McElwee J, Podtelezhnikov AA, et al. Integrated systems approach identifies genetic nodes and networks in late-onset Alzheimer’s disease. Cell. 2013;153:707–20.

39. Lanke V, Moolamalla STR, Roy D, Vinod PK. Integrative Analysis of Hippocampus Gene Expression Profiles Identifies Network Alterations in Aging and Alzheimer’s Disease. Front Aging Neurosci. 2018;10:153.

40. De Strooper B, Scorrano L. Close encounter: mitochondria, endoplasmic reticulum and Alzheimer’s disease. EMBO J. 2012;31:4095–7.

41. Hashimoto S, Saido TC. Critical review: involvement of endoplasmic reticulum stress in the aetiology of Alzheimer’s disease. Open Biol. 2018;8.

42. Area-Gomez E, de Groof AJ, Boldogh I, Bird TD, Gibson GE, Koehler CM, et al. Presenilins are enriched in endoplasmic reticulum membranes associated with mitochondria. Am J Pathol. 2009;175:1810–6.

43. Baloyannis SJ. Mitochondria are related to synaptic pathology in Alzheimer’s disease. Int J Alzheimers Dis. 2011;2011:305395.

44. Horvath S. DNA methylation age of human tissues and cell types. Genome Biol. 2013;14:R115.

45. Grodstein F, Lemos B, Yu L, Klein HU, Iatrou A, Buchman AS, et al. The association of epigenetic clocks in brain tissue with brain pathologies and common aging phenotypes. Neurobiol Dis. 2021;157:105428.

