## Supplementary Materials and Methods for "Analysis of Alzheimer’s disease Polygenic Risk Scores using RNA-sequencing provides further novel biological pathways"

##### Suppl. Table 1. Dataset description and numbers

a) MayoRNAseq sample description

| Tissue | N WGS samples | N RNA-seq samples | N matched RNA-seq WGS* | N genetically unique* | Diagnosis* | mean age at death* | Sex* |
|---|---|---|---|---|---|---|---|
| cerebellum | 159 | 159 | 144 | 144 | 76 (AD)<br>68 (CON) | 82.7 (AD)<br>82.2 (CON) | 66 male<br>78 female |
| temporal cortex | 160 | 160 | 144 | 144 | 77 (AD)<br>67 (CON) | 82.8 (AD)<br>82.5 (CON) | 65 male<br>79 female |
| **Total** | **319** | **319** | **288** | **170** | **153 (AD)<br>135 (CON)** | **82.8 (AD)<br>82.3 (CON)** | **131 (male)<br>157 (female)** |

*after QC

##### Suppl. Table 2. RNA-SeQC measures used to filter out RNA-seq individuals

| QC measure | cut-off threshold | derived from |
|---|---|---|
| Mapping Rate | <4x sd | RNA-SeQC |
| Duplicate Rate of Mapped | >4x sd | RNA-SeQC |
| Duplicate Rate of Mapped excluding Globins | >4x sd | RNA-SeQC |
| Expression Profiling Efficiency | <4x sd | RNA-SeQC |
| High Quality Rate | <4x sd | RNA-SeQC |
| Exonic Rate | <4x sd | RNA-SeQC |
| Intronic Rate | >4x sd | RNA-SeQC |
| Intergenic Rate | >4x sd | RNA-SeQC |
| Intragenic Rate | <4x sd | RNA-SeQC |
| Ambiguous Alignment Rate | >4x sd | RNA-SeQC |
| High Quality Exonic Rate | <4x sd | RNA-SeQC |
| High Quality Intronic Rate | >4x sd | RNA-SeQC |
| High Quality Intergenic Rate | >4x sd | RNA-SeQC |
| High Quality Intragenic Rate | <4x sd | RNA-SeQC |
| High Quality Ambiguous Alignment Rate | >4x sd | RNA-SeQC |
| rRNA Rate | >4x sd | RNA-SeQC |
| End 1 Sense Rate | >4x sd | RNA-SeQC |
| End 2 Sense Rate | <4x sd | RNA-SeQC |
| Genes Detected | <15,000 and >30,000 | RNA-SeQC |
| Median 3' bias | >4x sd | RNA-SeQC |
| 3' bias Std | >4x sd | RNA-SeQC |
| 3' bias MAD Std | >4x sd | RNA-SeQC |
| 3' Bias, 25th Percentile | >4x sd | RNA-SeQC |
| 3' Bias, 75th Percentile | >4x sd | RNA-SeQC |
| Median of Transcript Coverage CV | >4x sd | RNA-SeQC |
| Median Exon CV | >4x sd | RNA-SeQC |
| Exon CV MAD | >4x sd | RNA-SeQC |
| Chimeric Reads Rate | >1e-05 | $\frac{\text{Chimeric reads}}{\text{Total Mapped Reads}}$ |

| | | |
|---|---|---|
| End 1 Antisense Rate | <4x sd (depending on +/- strand) | $\frac{\text{End 1 Antisense}}{\text{Total Mapped Reads}}$ |
| End 2 Antisense Rate | >4x sd (depending on +/- strand) | $\frac{\text{End 2 Antisense}}{\text{Total Mapped Reads}}$ |
| Low Mapping Quality Rate | >4x sd | $\frac{\text{Low Mapping Quality}}{\text{Total Mapped Reads}}$ |
| Low Quality Reads Rate | >4x sd | $\frac{\text{Low Quality Reads}}{\text{Total Mapped Reads}}$ |
| Non-Globin Reads Rate | <0.9 | $\frac{\text{Alternative Alignments}}{\text{Total Mapped Reads}}$ |
| Non-Globin Duplicate Reads Rate | >4x sd | $\frac{\text{Non} - \text{Globin Duplicate Reads}}{\text{Total Mapped Reads}}$ |
| Unique Mapping, Vendor QC Passed Reads Rate | <0.5 | $\frac{\text{Unique Mapping, Vendor QC Passed Reads}}{\text{Total Mapped Reads}}$ |

#### Suppl. Table3. Assigning *APOE* status

a) *APOE* alleles

| rs429358 | rs7412 | *APOE* name |
|---|---|---|
| C | T | ε1 |
| T | T | ε2 |
| T | C | ε3 |
| C | C | ε4 |

b) Coding used for the *APOE* haplotypes

| *APOE* name | rs429358 | rs7412 | Coding used | Comment |
|---|---|---|---|---|
| ε1/ε1 | (C;C) | (T;T) | 11 | |
| ε1/ε2 | (C;T) | (T;T) | 12 | |
| ε2/ε4 or ε1/ε3 | (C;T) | (C;T) | 24 | ambiguous, ε2/ε4 or ε1/ε3 |
| ε2/ε4 or ε1/ε3 | (C;T) | (C;T) | 24 | ambiguous, ε2/ε4 or ε1/ε3 |
| ε1/ε4 | (C;C) | (C;T) | 14 | |
| ε2/ε2 | (T;T) | (T;T) | 22 | |
| ε2/ε3 | (T;T) | (C;T) | 23 | |
| ε3/ε3 | (T;T) | (C;C) | 33 | |
| ε3/ε4 | (C;T) | (C;C) | 34 | |
| ε4/ε4 | (C;C) | (C;C) | 44 | |

**Suppl. Figure 1. MayoRNAseq WGS PCAs (with 1,000 genomes phase3) and rna-seq PCA (CQN-normalised gene-expression)**

a) MayoRNAseq rna-seq PCA (cerebellum), CQN-normalised gene counts

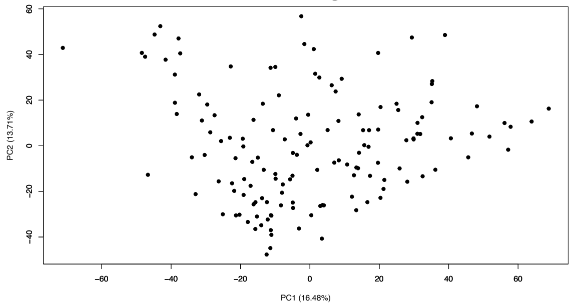


b) MayoRNAseq rna-seq PCA (temporal cortex), CQN-normalised gene counts

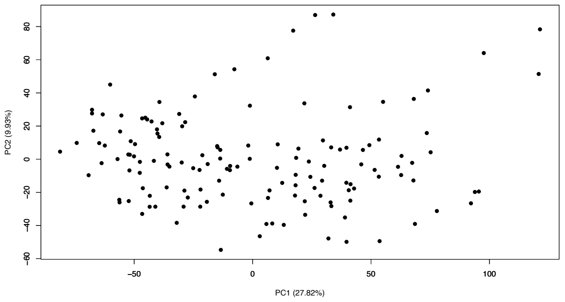


c) MayoRNAseq WGS PCA with 1,000 genomes after QC

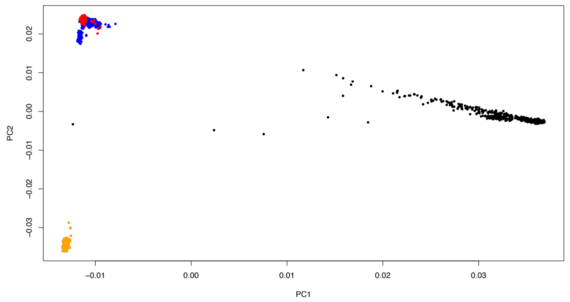


Red dots MayoRNAseq samples; Black dots 1,000 genomes AFR; Orange dots 1,000 genomes ASN; Blue dots 1,000 genomes EUR

#### Suppl. Figure 2. Overlap of DE genes in cerebellum and temporal cortex tissues in MayoRNAseq (case/control with *APOE* status)

a) rank plot of all genes. X-axis cerebellum, Y-axis temporal cortex. p=7.40e-88, $r^2$=0.018

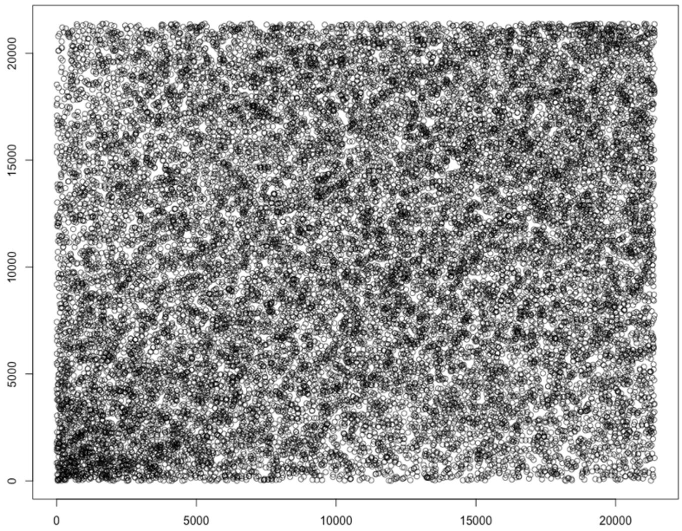


b) diff. vs. diff (p=2.22e-64)

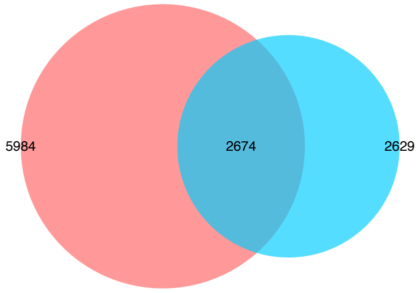


c) up vs. up (p=7.18e-138)

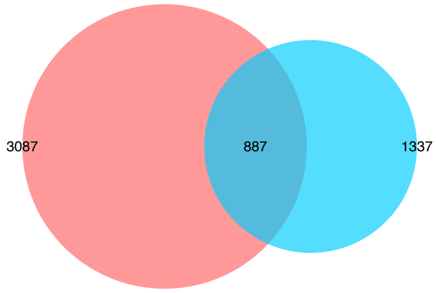


d) down vs. down (p<1e-300)

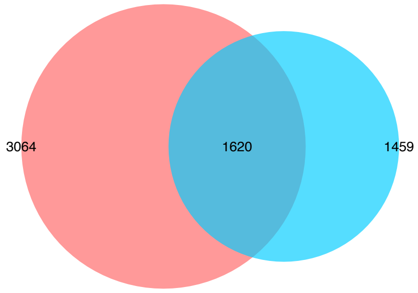


e) up vs. down (p=1)

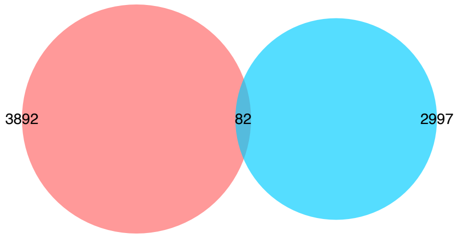


f) down vs. up (p=1)

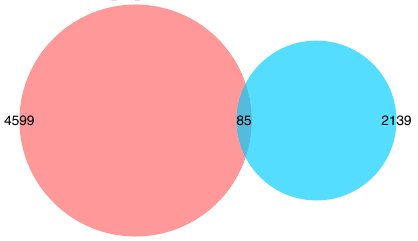


*Proportional Venn diagram. Numbers represent the significant DE genes (FDR) in the two lists with the middle number representing the number of genes that overlap. Red colour represents cerebellum the green temporal cortex. p-values derived from hypergeometric test. up and down represent up-regulated and down-regulated genes respectively. a) most significant gene has rank of 1.

#### Suppl. Figure 3. Overlap of GO terms in cerebellum and temporal cortex tissues in MayoRNAseq (case/control with *APOE* status)

a) no-direction (p=7.78e-11)

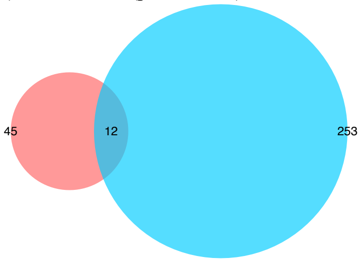


b) up vs. up (p=4.11e-287)

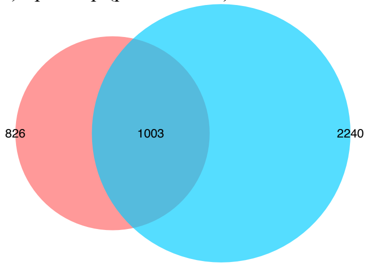


c) down vs. down (p=7.93e-85)

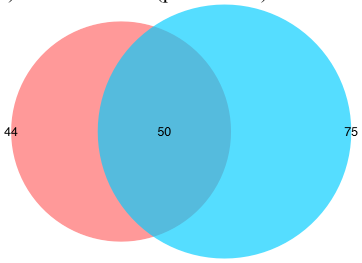


d) up vs. down (p=9.95e-01)

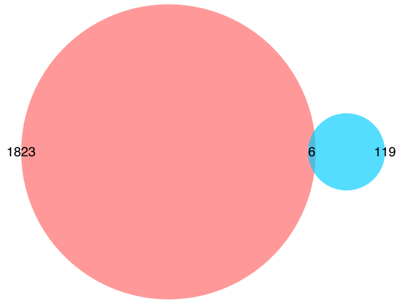


e) down vs. up (p=1)

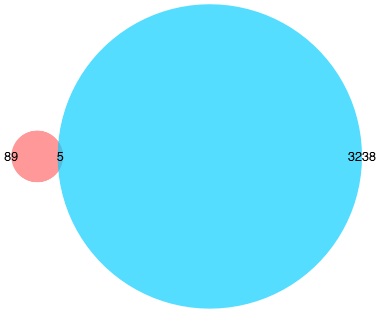


f) rank plot no-direction GO terms. x-axis cerebellum, y-axis temporal cortex. p=1.76e-04, r2=0.000845

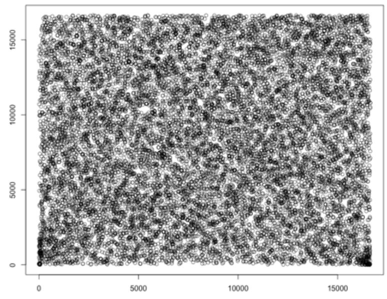


g) rank plot up-regulated GO terms. x-axis cerebellum, y-axis temporal cortex. p<2e-16, r2=0.21

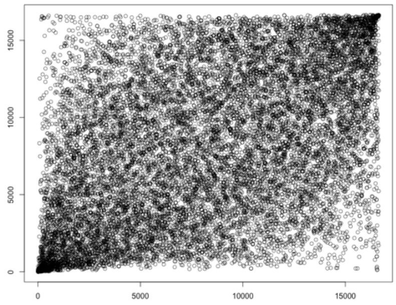


h) rank plot down-regulated GO terms. x-axis cerebellum, y-axis temporal cortex. p<2e-16, r2=0.21

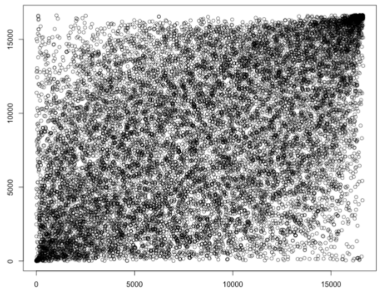


*Proportional Venn diagram. Numbers represent the significant GO terms (FDR) in the two lists with the middle number representing the number of genes that overlap. Red colour represents cerebellum the blue temporal cortex. p-values derived from hypergeometric test. f-h, most significant GO term has rank of 1

**Suppl. Figure 4. GO term semantic similarity clustering, case/control cerebellum and case/control temporal cortex (gene order based on p-values only; GO no direction)**

a) Biological Process (BP)

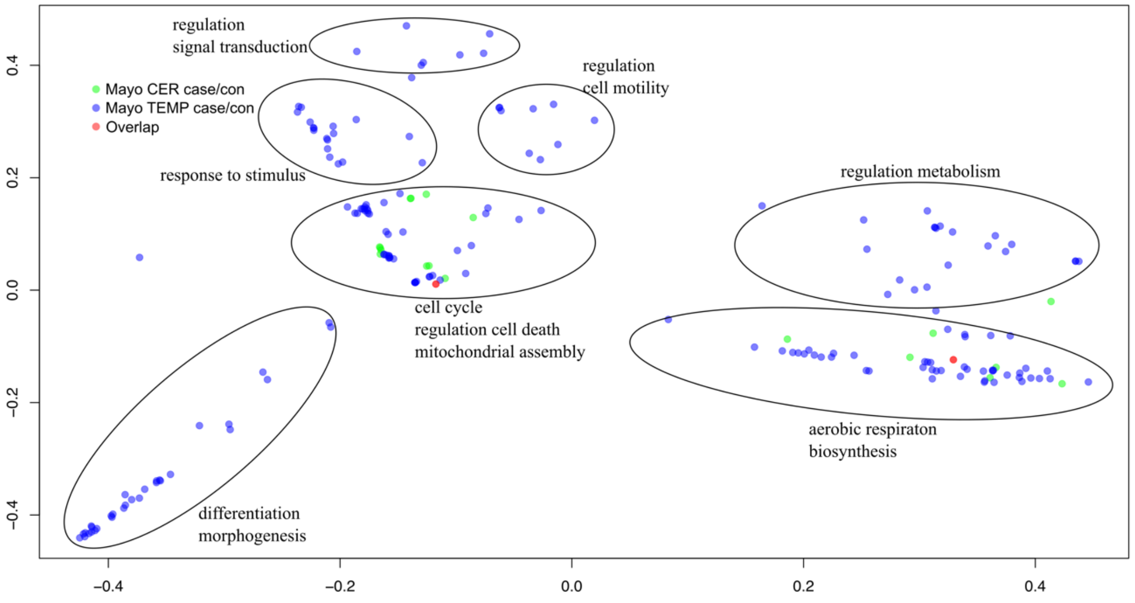


b) Cellular Component (CC)

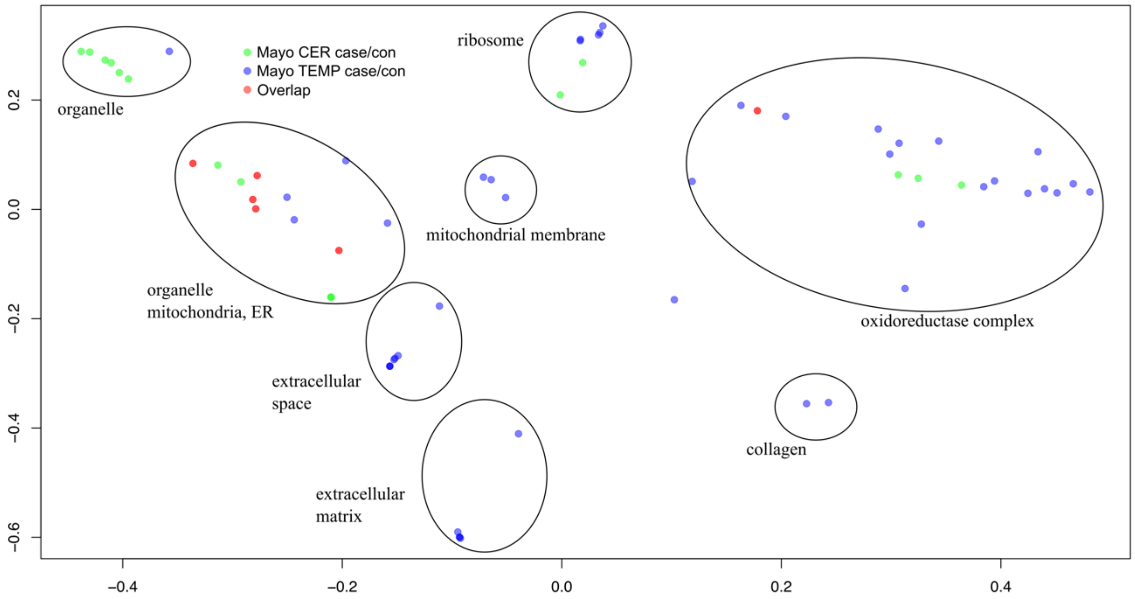


X and Y axes represent CMD dimension 1 and 2. GO term p≤0.05 FDR. Green dots represent significant GO terms from the case/control analysis of cerebellum, Blue dots represent significant GO terms from the case/control analysis of temporal cortex, Red dots represent significant GO terms overlapping in case/control analysis of cerebellum and temporal cortex. Cluster labels were manually curated based on the most common GO term in the cluster.

**Suppl. Figure 5. GO term semantic similarity clustering, case/control cerebellum and case/control temporal cortex (gene order most up-regulated at top; GO up-regulated)**

a) Biological Process (BP)

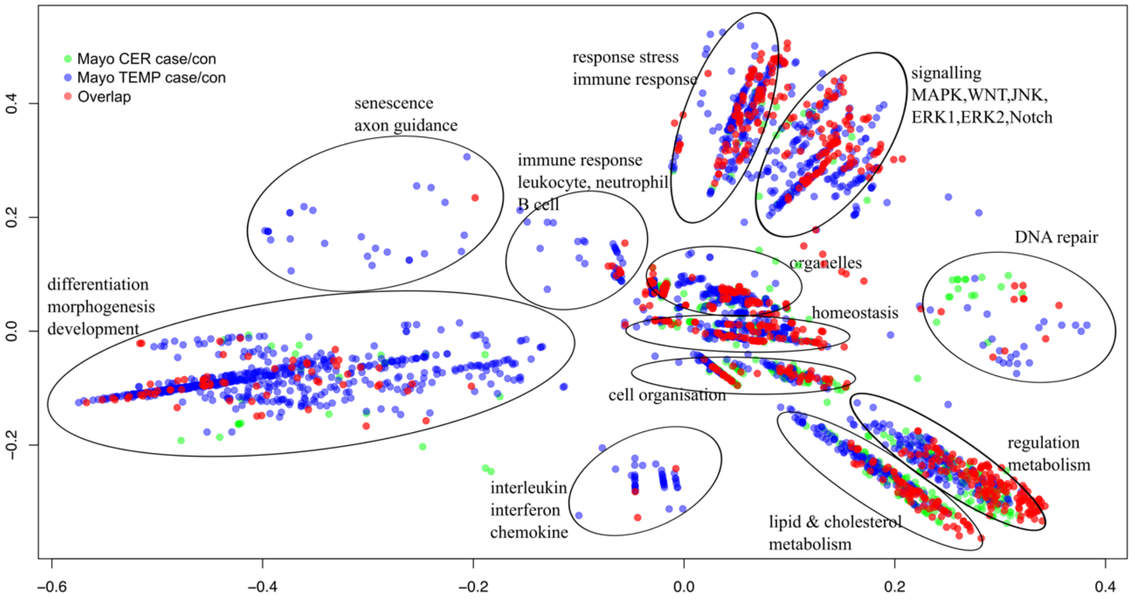


b) Cellular Component (CC)

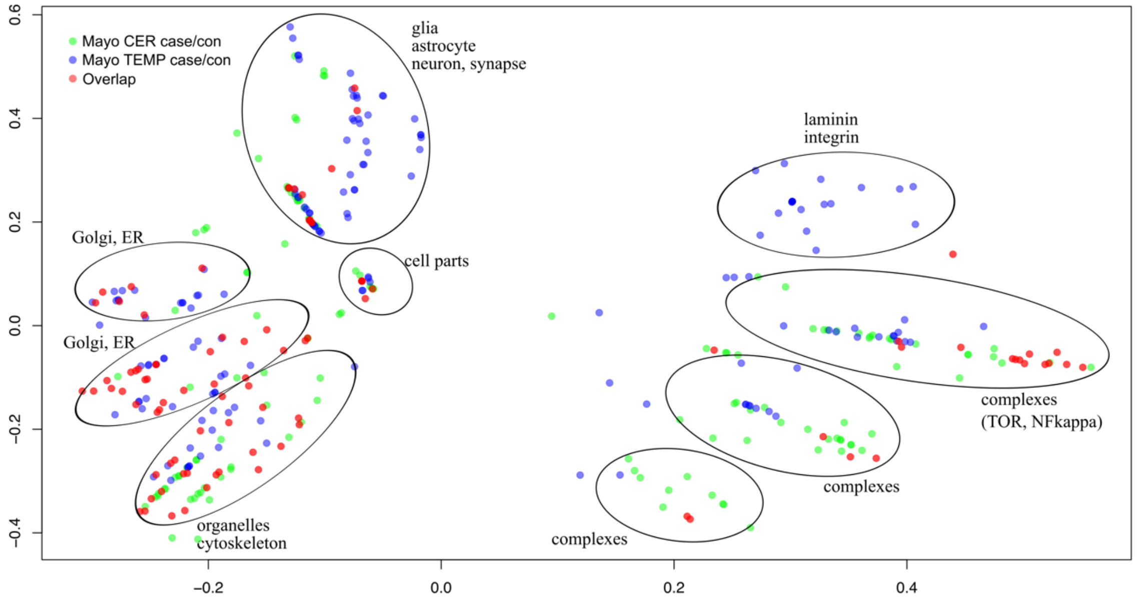


X and Y axes represent CMD dimension 1 and 2. GO term p≤0.05 FDR. Green dots represent significant GO terms from the case/control analysis of cerebellum, Blue dots represent significant GO terms from the case/control analysis of temporal cortex, Red dots represent significant GO terms overlapping in case/control analysis of cerebellum and temporal cortex. Cluster labels were manually curated based on the most common GO term in the cluster.

**Suppl. Figure 6. GO term semantic similarity clustering, case/control cerebellum and case/control temporal cortex (gene order most down-regulated at top; GO down-regulated)**

a) Biological Process (BP)

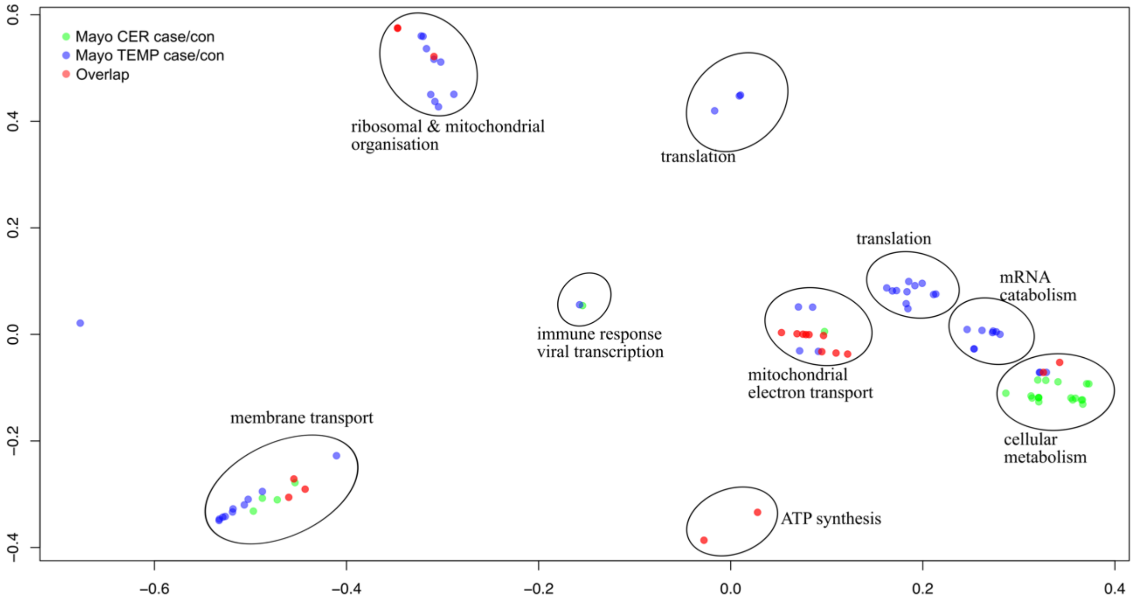


b) Cellular Component (CC)

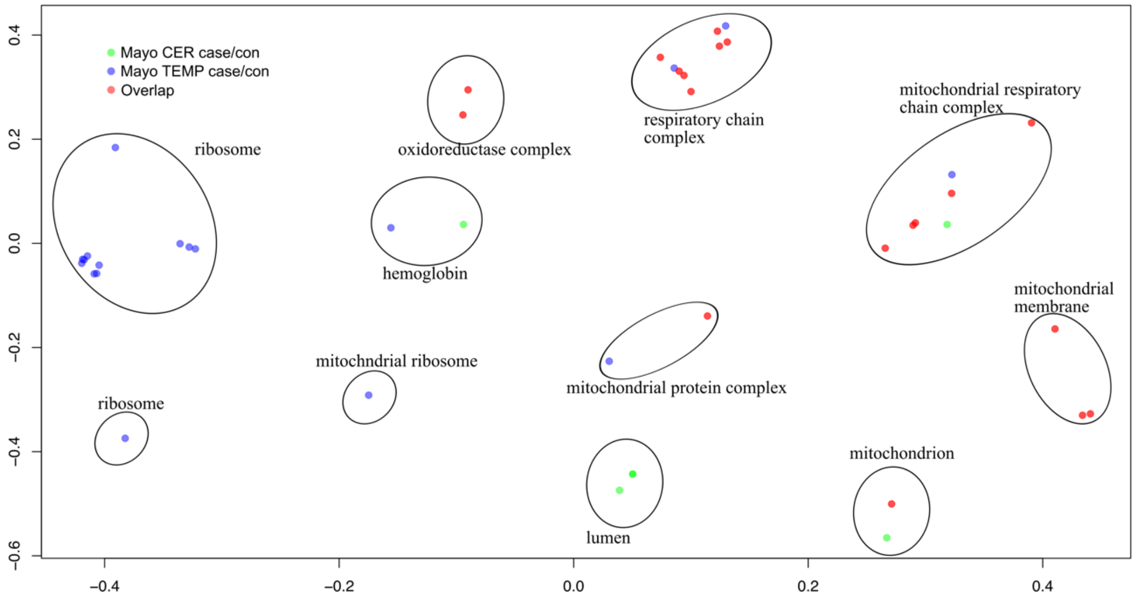


X and Y axes represent CMD dimension 1 and 2. GO term p≤0.05 FDR. Green dots represent significant GO terms from the case/control analysis of cerebellum, Blue dots represent significant GO terms from the case/control analysis of temporal cortex, Red dots represent significant GO terms overlapping in case/control analysis of cerebellum and temporal cortex. Cluster labels were manually curated based on the most common GO term in the cluster.

**Suppl. Figure 7. GO term enrichment comparison Catmap vs. topGO, case/control cerebellum and case/control temporal cortex**

Cerebellum

a) no-direction vs. no-direction
(p-value<1e-16; $r^2$=0.74)

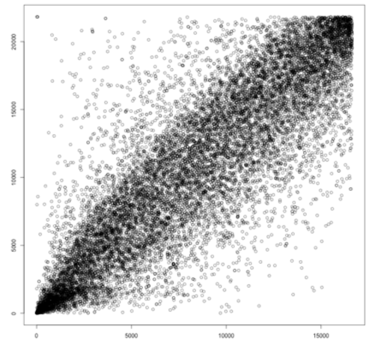

c) up-regulated vs. up-regulated
(p-value<1e-16; r2=0.79)

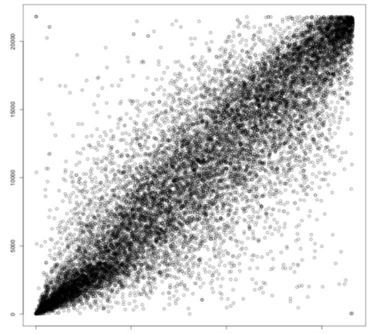

e) down-regulated vs. down-regulated
(p-value<1e-16; r2=0.65)

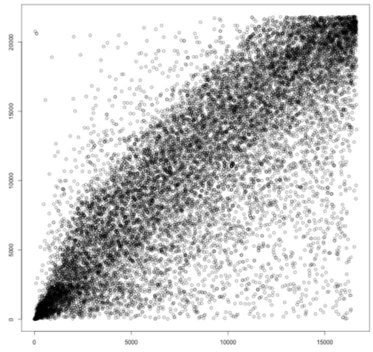

Temporal cortex

b) no-direction vs. no-direction
(p-value<1e-16; r2=0.73)

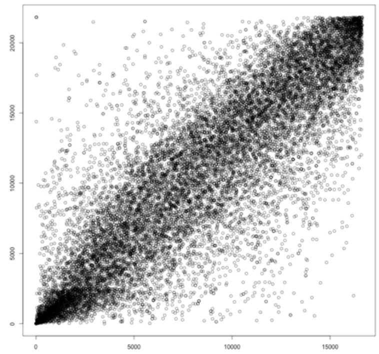

d) up-regulated vs. up-regulated
(p-value<1e-16; r2=0.80)

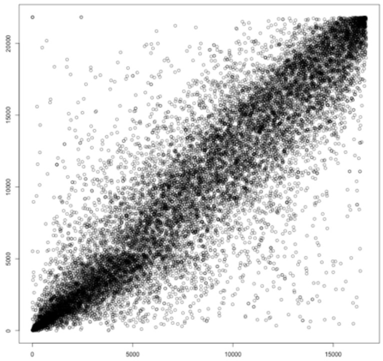

f) down-regulated vs. down-regulated
(p-value<1e-16; r2=0.69)

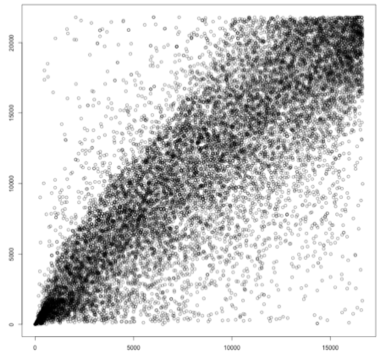

X-axis represents ranks of GO-terms derived by Catmap and Y-axis represent GO-terms derived by topGO (classic algorithm with ks statistic or Kolmogorov-Smirnov test). p-values and $r^2$ were derived using a linear model. The most significant GO term (p-value) will have a rank of 1.

#### Suppl. Figure 8. Overlap of top 300 DE genes in cerebellum and temporal cortex tissues in MayoRNAseq (PRS with *APOE* status)

a) rank plot of all genes. X-axis cerebellum, Y-axis temporal cortex. p=3.34e-80, $r^2$=0.017

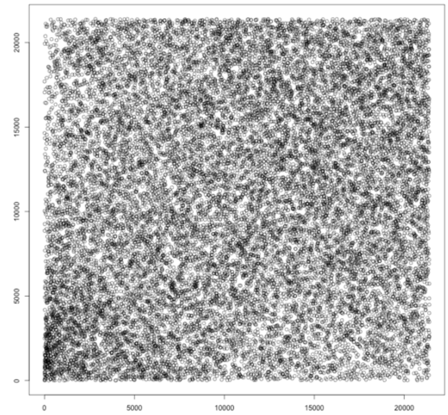


b) diff. vs. diff (p=3.57e-03)

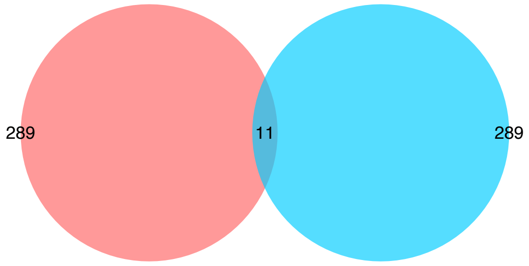


a) up vs. up (p=5.51e-06)

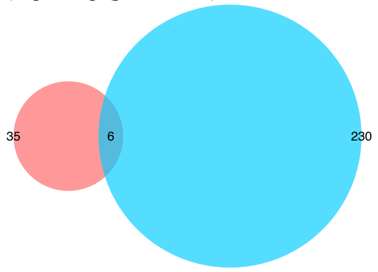


b) down vs. down (p=1.07e-03)

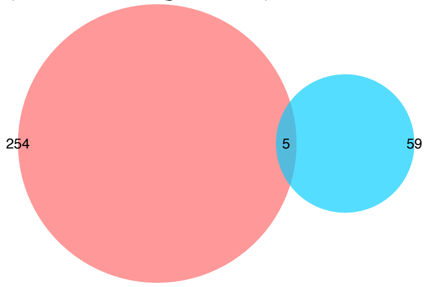


a) up vs. down (p=1)

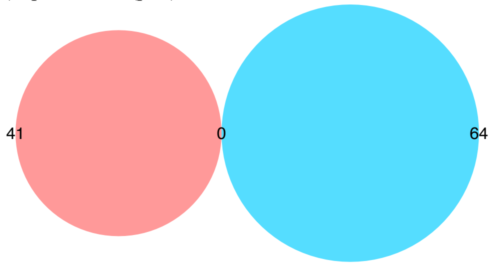


b) down vs. up (p=1)

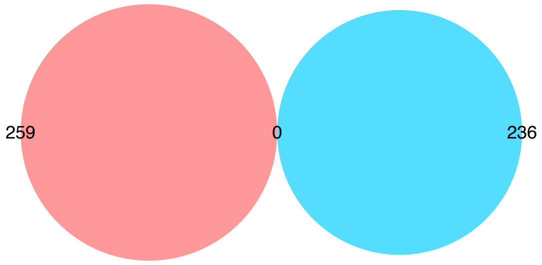


*Proportional Venn diagram. Numbers represent the significant DE genes (FDR) in the two lists with the middle number representing the number of genes that overlap. Red colour represents cerebellum the green temporal cortex. p-values derived from hypergeometric test. up and down represent up-regulated and down-regulated genes respectively. a) most significant gene has rank of 1.

#### Suppl. Figure 9. Overlap of GO terms in cerebellum and temporal cortex tissues in MayoRNAseq (PRS with *APOE* status)

a) no-direction (p=9.40e-14)

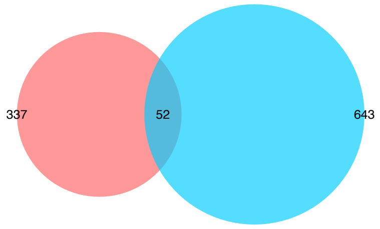


b) up vs. up (p<1e-300)

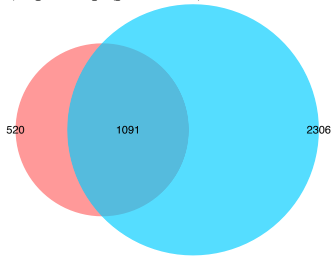


c) down vs. down (p=6.85e-65)

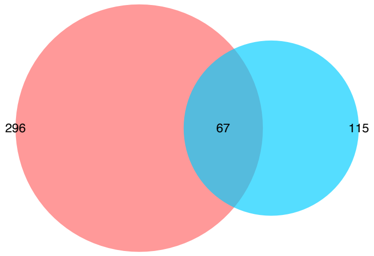


d) up vs. down (p=3.09e-01)

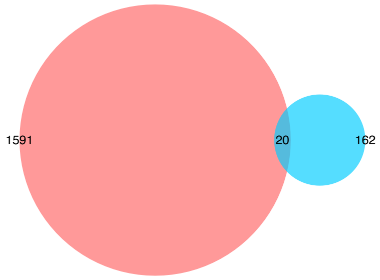


e) down vs. up (p=5.34e-02)

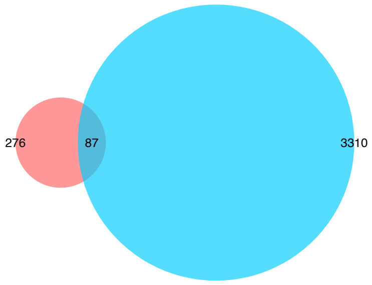


f) rank plot no-direction GO terms. x-axis cerebellum, y-axis temporal cortex. p<2e-16, r2=0.018

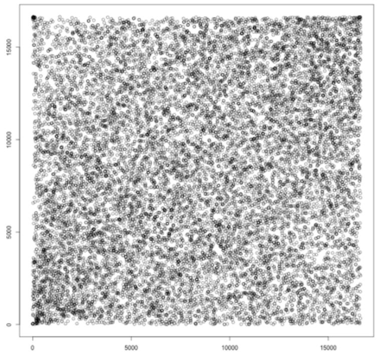

g) rank plot up-regulated GO terms. x-axis cerebellum, y-axis temporal cortex. p<2e-16, r2=0.21

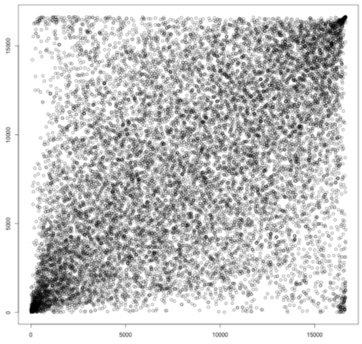

h) rank plot down-regulated GO terms. x-axis cerebellum, y-axis temporal cortex. p<2e-16, r2=0.21

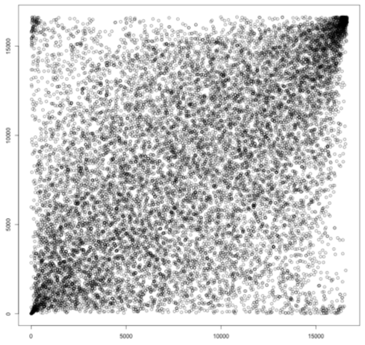

*Proportional Venn diagram. Numbers represent the significant GO terms (FDR) in the two lists with the middle number representing the number of genes that overlap. Red colour represents cerebellum the blue temporal cortex. p-values derived from hypergeometric test. f-h, most significant GO term has rank of 1

**Suppl. Figure 10. GO term semantic similarity clustering, PRS cerebellum and PRS temporal cortex (gene order based on p-values only; GO no direction)**

a) Biological Process (BP)

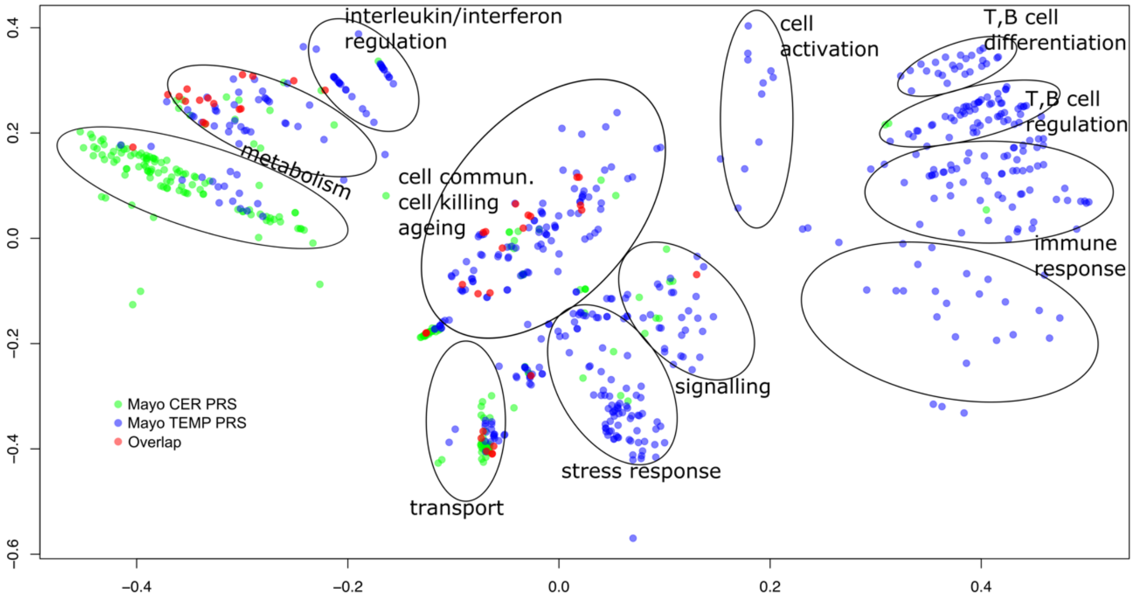


b) Cellular Component (CC)

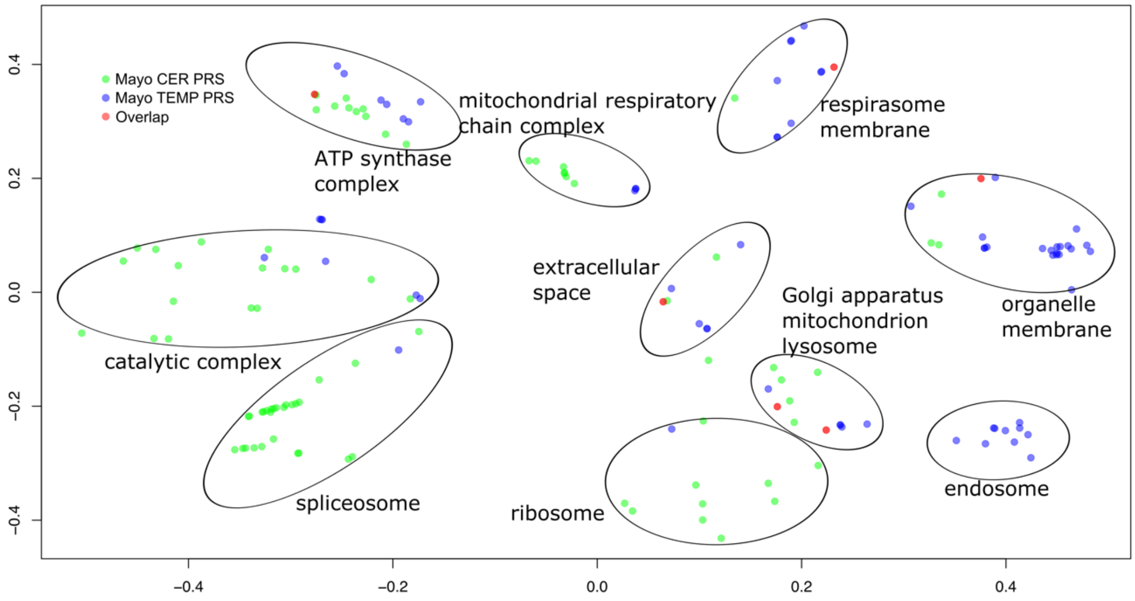


X and Y axes represent CMD dimension 1 and 2. GO term p≤0.05 FDR. Green dots represent significant GO terms from the PRS analysis of cerebellum, Blue dots represent significant GO terms from the PRS analysis of temporal cortex, Red dots represent significant GO terms overlapping in PRS analysis of cerebellum and temporal cortex. Cluster labels were manually curated based on the most common GO term in the cluster.

**Suppl. Figure 11. GO term semantic similarity clustering, PRS cerebellum and PRS temporal cortex (gene order most up-regulated at top; GO up-regulated)**

a) Biological Process (BP)

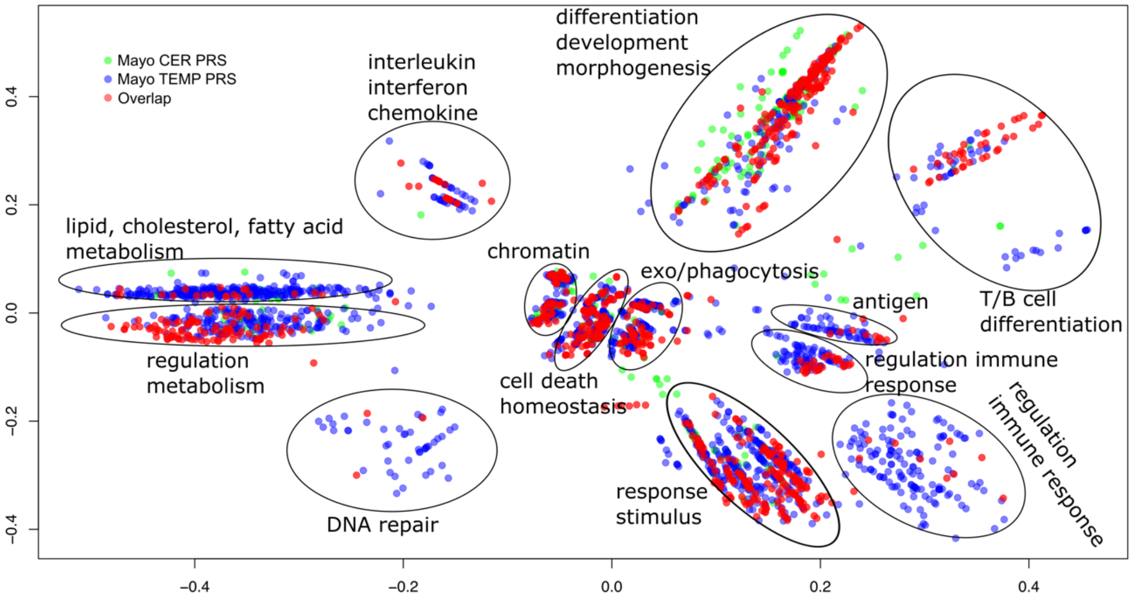


b) Cellular Component (CC)

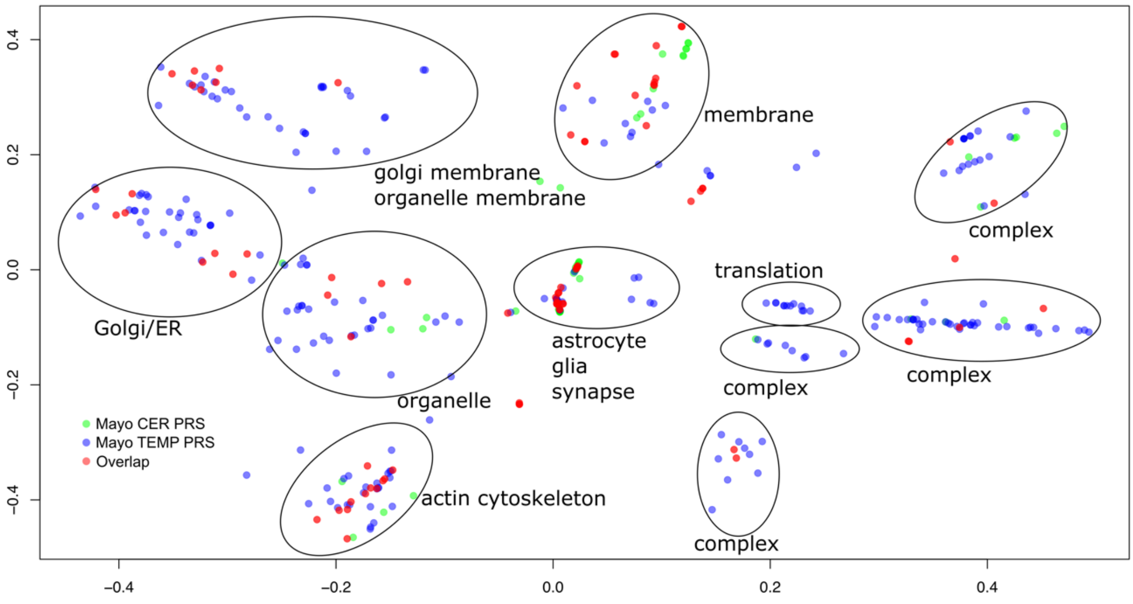


X and Y axes represent CMD dimension 1 and 2. GO term p≤0.05 FDR. Green dots represent significant GO terms from the PRS analysis of cerebellum, Blue dots represent significant GO terms from the PRS analysis of temporal cortex, Red dots represent significant GO terms overlapping in PRS analysis of cerebellum and temporal cortex. Cluster labels were manually curated based on the most common GO term in the cluster.

**Suppl. Figure 12. GO term semantic similarity clustering, PRS cerebellum and PRS temporal cortex (gene order most down-regulated at top; GO down-regulated)**

a) Biological Process (BP)

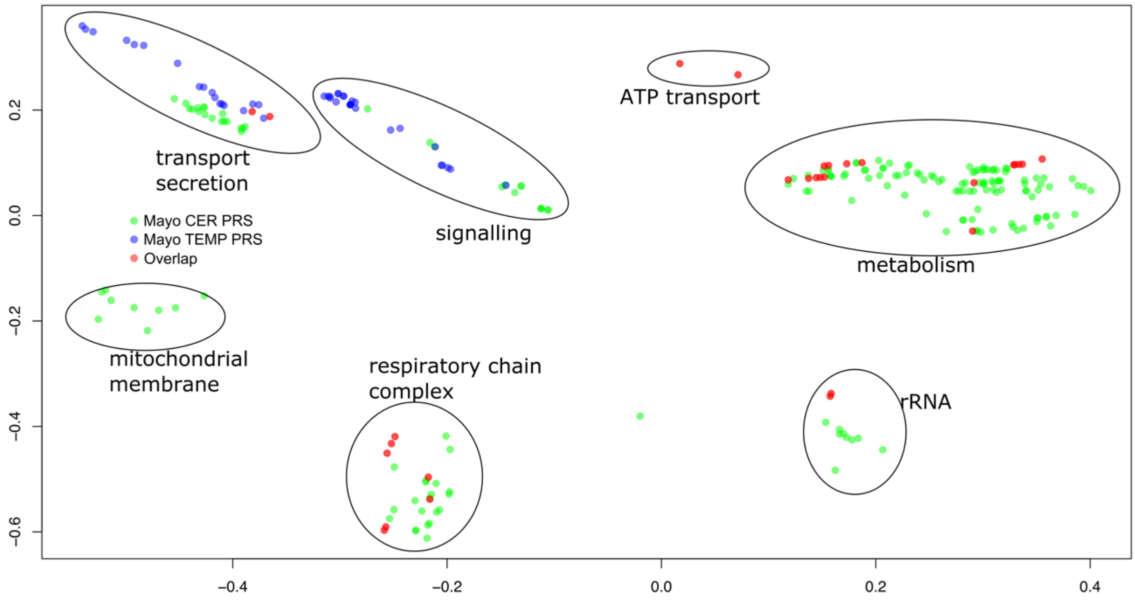


b) Cellular Component (CC)

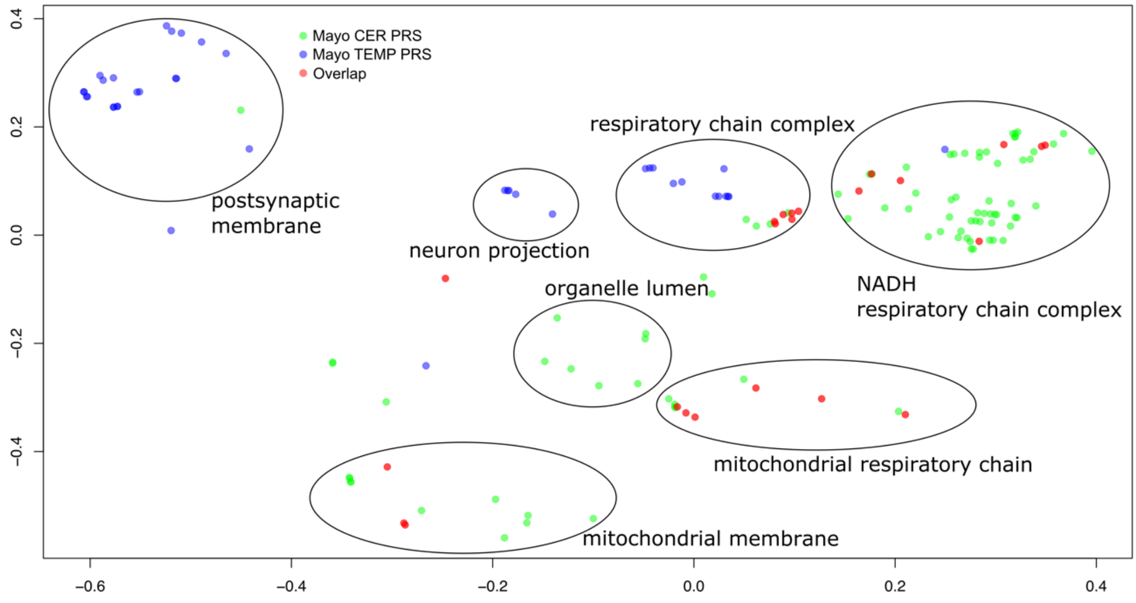


X and Y axes represent CMD dimension 1 and 2. GO term p≤0.05 FDR. Green dots represent significant GO terms from the PRS analysis of cerebellum, Blue dots represent significant GO terms from the PRS analysis of temporal cortex, Red dots represent significant GO terms overlapping in PRS analysis of cerebellum and temporal cortex. Cluster labels were manually curated based on the most common GO term in the cluster.

**Suppl. Figure 13. GO term enrichment comparison Catmap vs. topGO, PRS cerebellum and PRS temporal cortex**

Cerebellum | Temporal cortex

a) no-direction vs. no-direction
(p-value<1e-16; $r^2$=0.68)

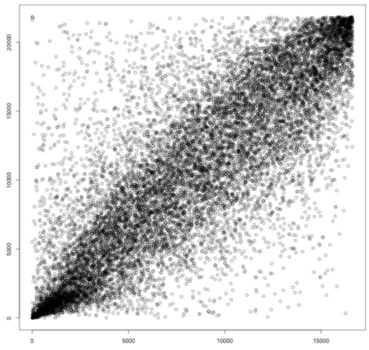

b) no-direction vs. no-direction
(p-value<1e-16; r2=0.75)

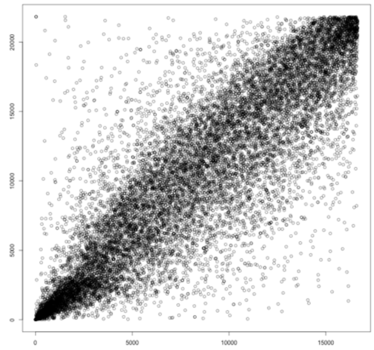

c) up-regulated vs. up-regulated
(p-value<1e-16; r2=0.81)

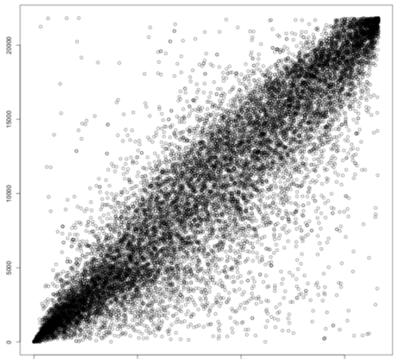

d) up-regulated vs. up-regulated
(p-value<1e-16; r2=0.79)

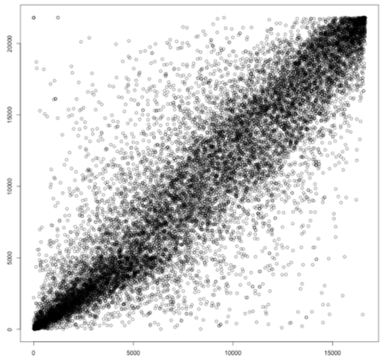

e) down-regulated vs. down-regulated
(p-value<1e-16; r2=0.76)

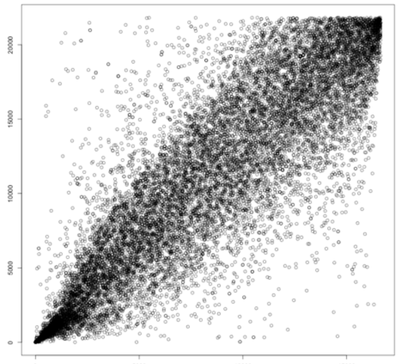

f) down-regulated vs. down-regulated
(p-value<1e-16; r2=0.65)

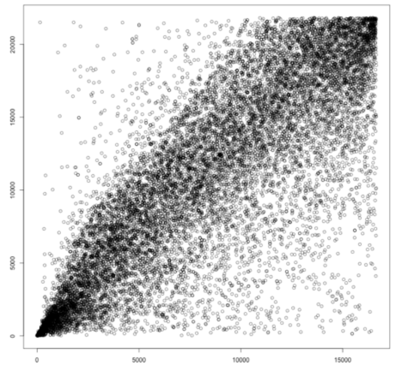

X-axis represents ranks of GO-terms derived by Catmap and Y-axis represent GO-terms derived by topGO (classic algorithm with ks statistic or Kolmogorov-Smirnov test). p-values and $r^2$ were derived using a linear model. The most significant GO term (p-value) will have a rank of 1.

#### Suppl. Figure 14. Overlap of DE genes MayoRNAseq (cerebellum case/control vs. PRS, both with *APOE* status)

a) rank plot of all genes. X-axis case/control cerebellum, Y-axis PRS cerebellum
$p<2.2e-16$, $r^2=0.03$

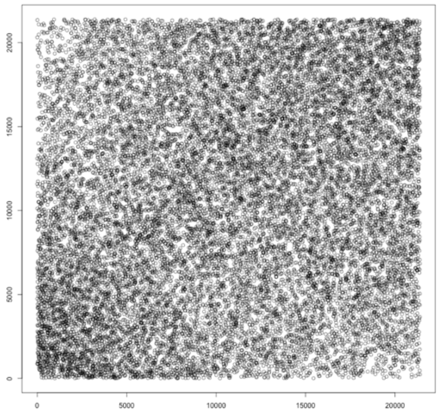


b) diff. vs. diff (p=7.89e-01)

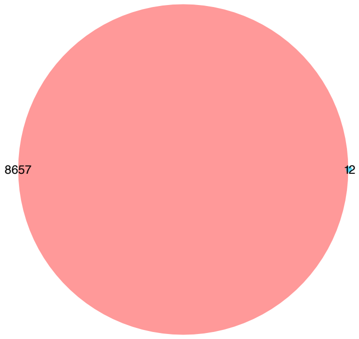


b) up vs. up (p=1)

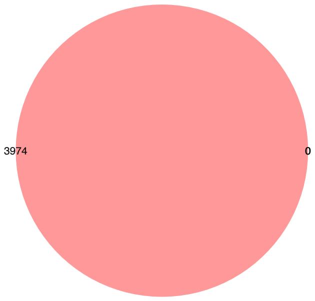


c) down vs. down (p=5.24e-01)

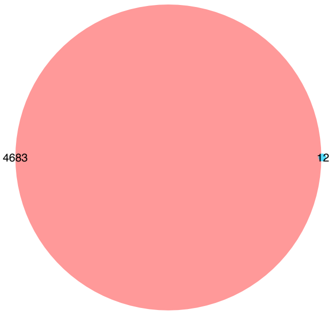


d) up vs. down (p=1)

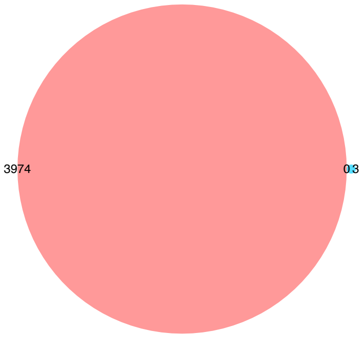


e) down vs. up (p=1)

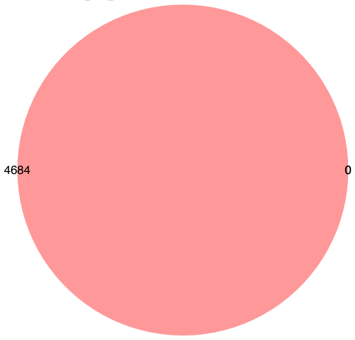


*Proportional Venn diagram. Numbers represent the significant DE genes (FDR) in the two lists with the middle number representing the number of genes that overlap. Red colour represents case/control the green full PRS. p-values derived from hypergeometric test. up and down represent up-regulated and down-regulated genes respectively.

**Suppl. Figure 15. Overlap of top 300 DE genes MayoRNAseq (cerebellum case/control vs. PRS, both with *APOE* status)**

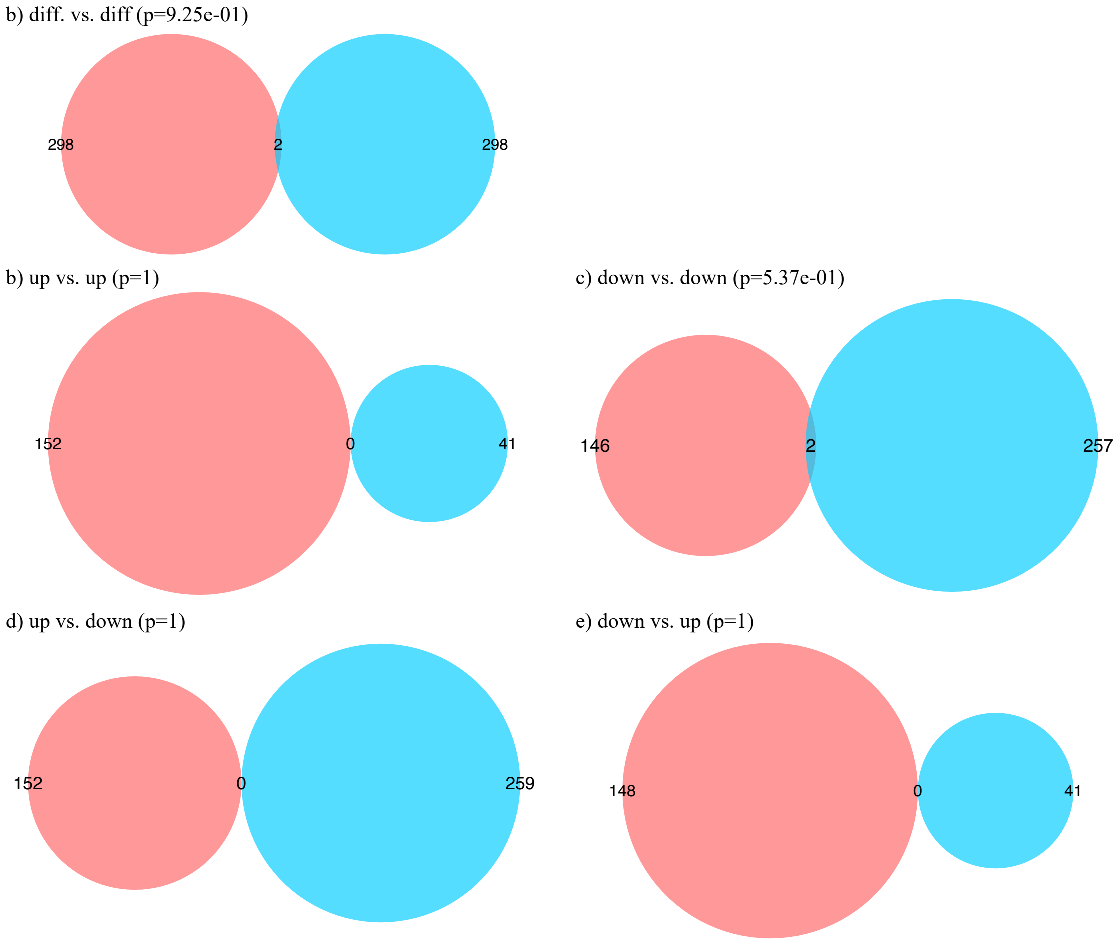


*Proportional Venn diagram. Numbers represent the significant DE genes (FDR) in the two lists with the middle number representing the number of genes that overlap. Red colour represents case/control the green full PRS. p-values derived from hypergeometric test. up and down represent up-regulated and down-regulated genes respectively.

#### Suppl. Figure 16. Overlap of DE genes MayoRNAseq (temporal cortex case/control vs. PRS, both with *APOE* status)

a) rank plot of all genes. X-axis case/control temporal cortex, Y-axis PRS temporal cortex
p=6.76e-04, $r^2$=0.00054

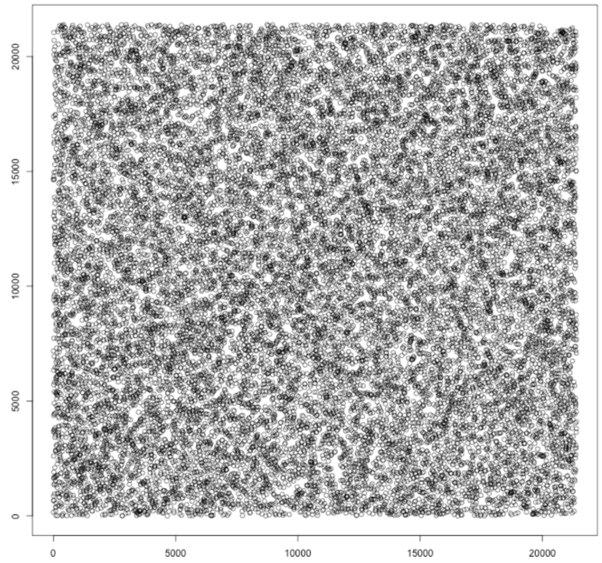


b) diff. vs. diff (p=2.86e-01)

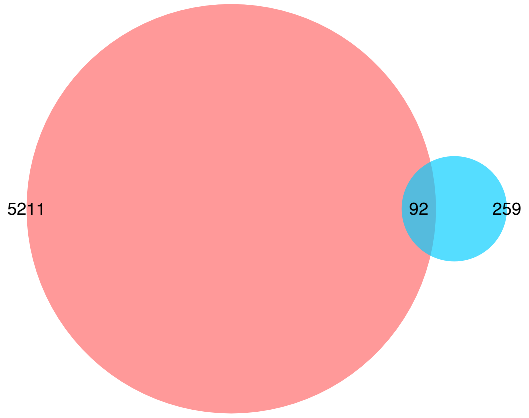


b) up vs. up (p=2.42e-11)

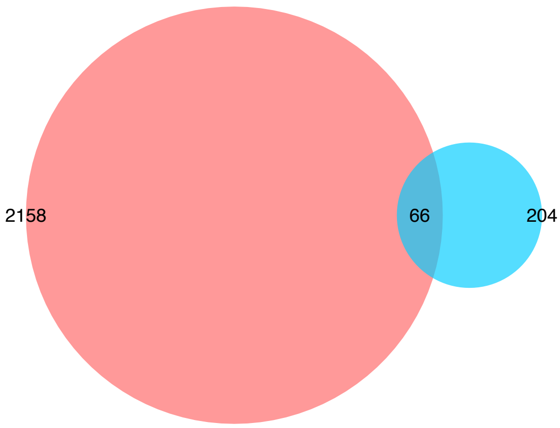


c) down vs. down (p=1.96e-03)

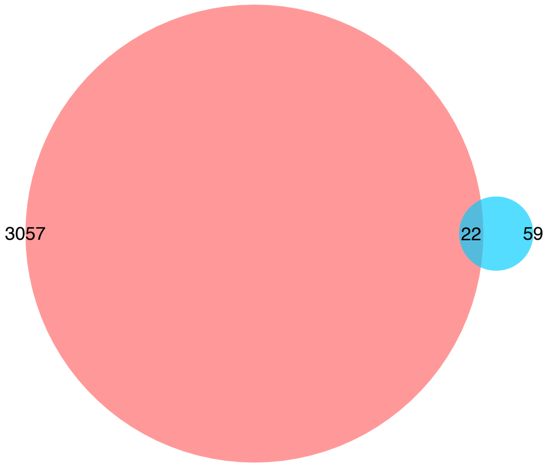


d) up vs. down (p=1)

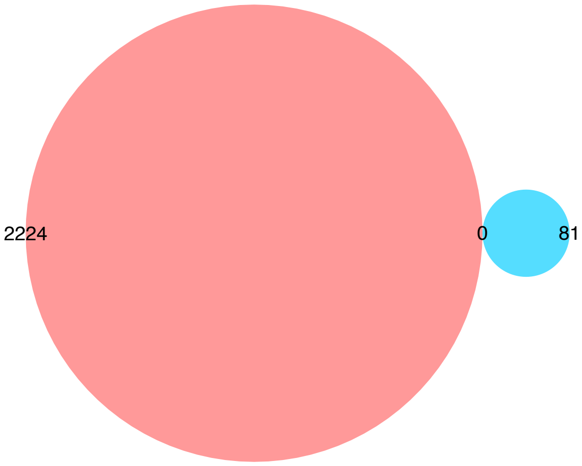


e) down vs. up (p=1)

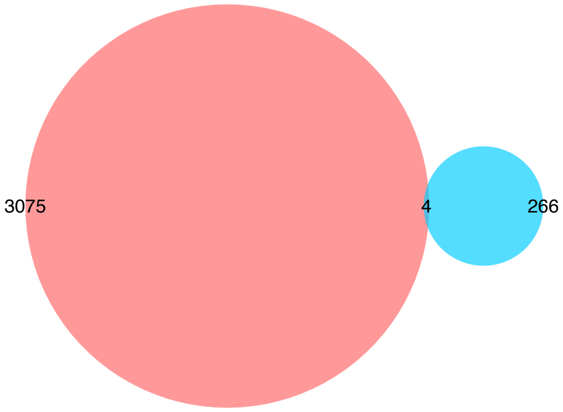


*Proportional Venn diagram. Numbers represent the significant DE genes (FDR) in the two lists with the middle number representing the number of genes that overlap. Red colour represents case/control the blue full PRS. p-values derived from hypergeometric test. up and down represent up-regulated and down-regulated genes respectively.

#### Suppl. Figure 17. Overlap of GO terms MayoRNAseq (cerebellum case/control vs. Full PRS, both with *APOE* status)

a) no-direction (p=4.09e-23)

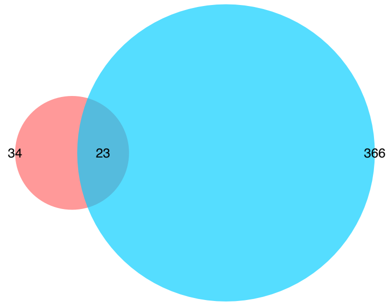


b) up vs. up (p=3.97e-243)

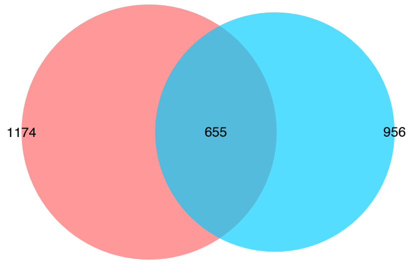


c) down vs. down (p=9.31e-66)

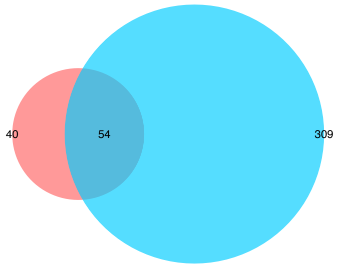


d) up vs. down (p=2.21e-03)

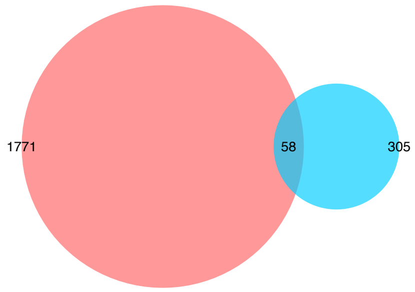


e) down vs. up (p=1)

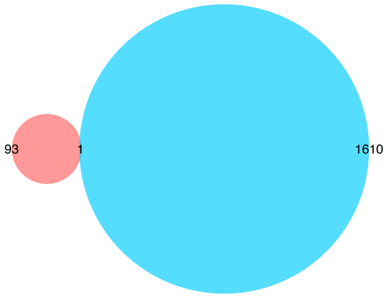


f) rank plot no-direction GO terms. x-axis cerebellum, y-axis temporal cortex. p<2e-16, r2=0.013

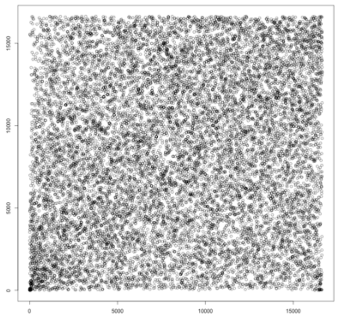

g) rank plot up-regulated GO terms. x-axis cerebellum, y-axis temporal cortex. p<2e-16, r2=0.22

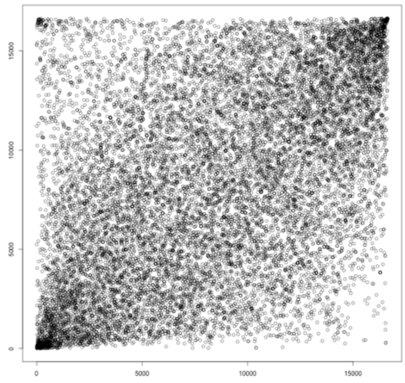

h) rank plot down-regulated GO terms. x-axis cerebellum, y-axis temporal cortex. p<2e-16, r2=0.22

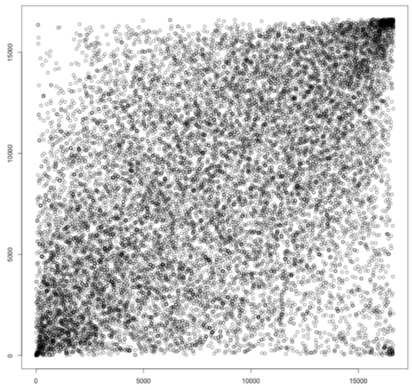

*Proportional Venn diagram. Numbers represent the significant GO terms (FDR) in the two lists with the middle number representing the number of genes that overlap. Red colour represents cerebellum the blue temporal cortex. p-values derived from hypergeometric test. f-h, most significant GO term has rank of 1

#### Suppl. Figure 18. Overlap of GO terms MayoRNAseq (temporal cortex case/control vs. Full PRS, both with *APOE* status)

a) no-direction (p=8.55e-19)

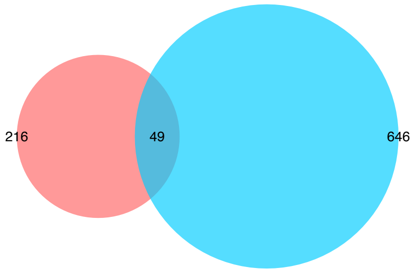


b) up vs. up (p<1e-300)

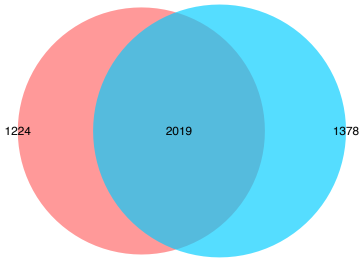


c) down vs. down (p=3.04e-93)

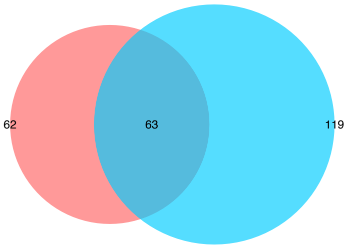


d) up vs. down (p=1)

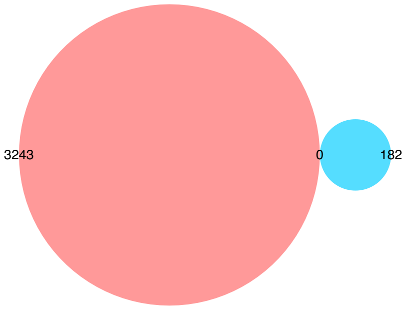


e) down vs. up (p=1.86e-01)

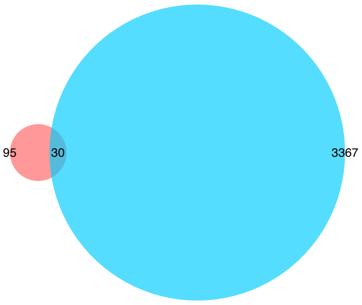


f) rank plot no-direction GO terms. x-axis cerebellum, y-axis temporal cortex. p<2e-16, r2=0.0047

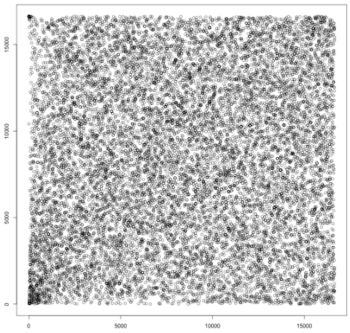

g) rank plot up-regulated GO terms. x-axis cerebellum, y-axis temporal cortex. p<2e-16, r2=0.39

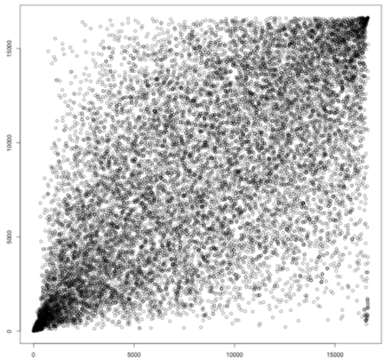


h) rank plot down-regulated GO terms. x-axis cerebellum, y-axis temporal cortex. p<2e-16, r2=0.39

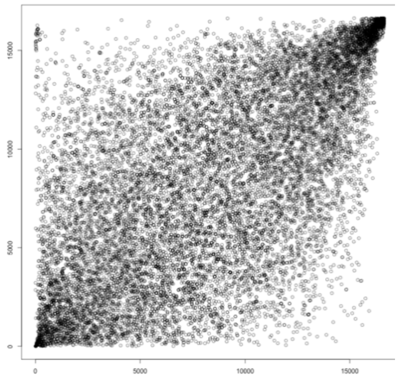


*Proportional Venn diagram. Numbers represent the significant GO terms (FDR) in the two lists with the middle number representing the number of genes that overlap. Red colour represents case/control temporal cortex the blue PRS temporal cortex. p-values derived from hypergeometric test. f-h, most significant GO term has rank of 1

**Suppl. Figure 19. GO term semantic similarity clustering, cerebellum case/control and PRS (gene order based on p-values only; GO no direction)**

a) Biological Process (BP)

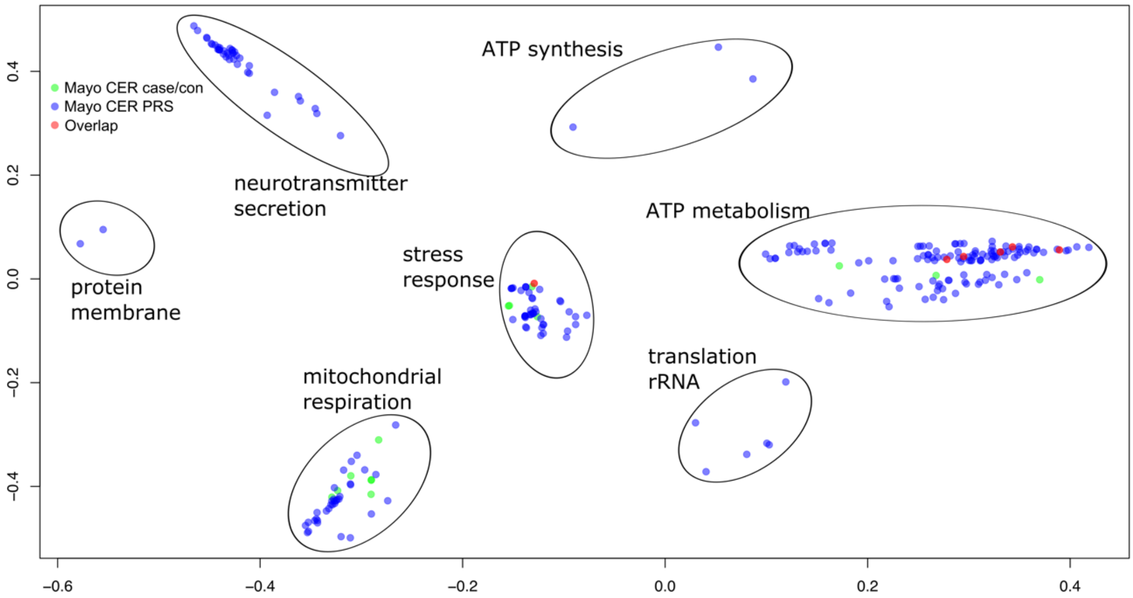


b) Cellular Component (CC)

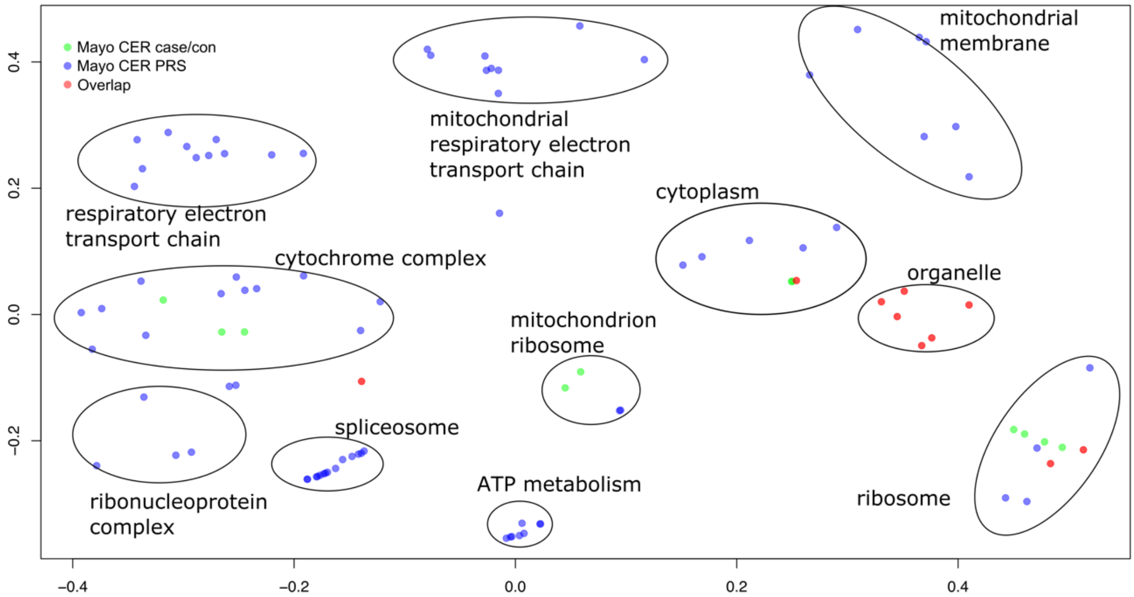


X and Y axes represent CMD dimension 1 and 2. GO term p≤0.05 FDR. Green dots represent significant GO terms from the PRS analysis of cerebellum, Blue dots represent significant GO terms from the PRS analysis of temporal cortex, Red dots represent significant GO terms overlapping in PRS analysis of cerebellum and temporal cortex. Cluster labels were manually curated based on the most common GO term in the cluster.

**Suppl. Figure 20. GO term semantic similarity clustering, cerebellum case/control and PRS (gene order most up-regulated at top; GO up-regulated)**

a) Biological Process (BP)

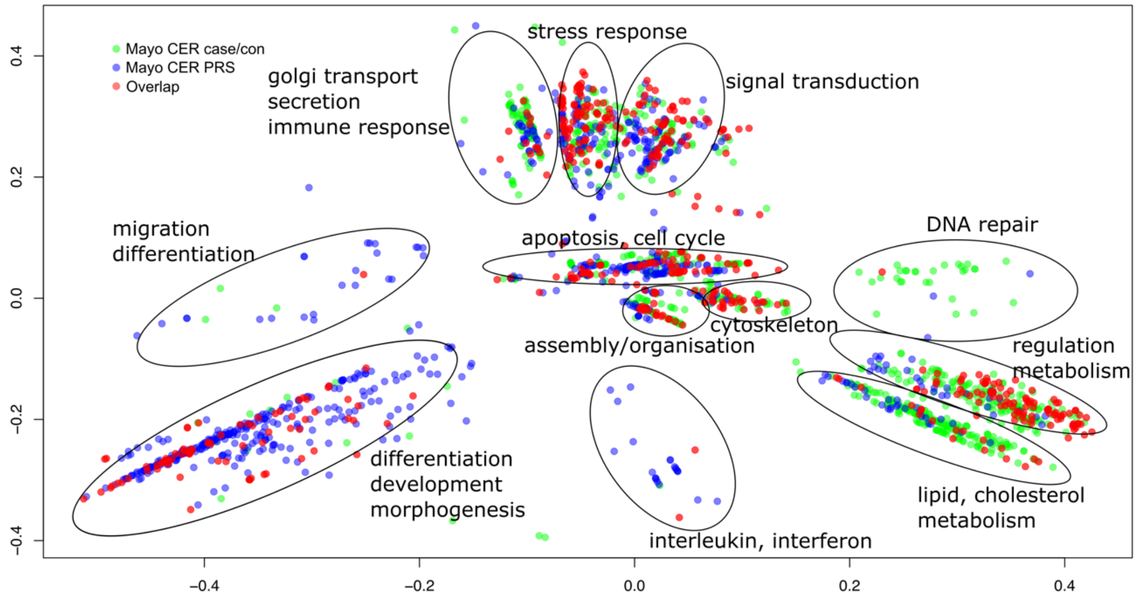


b) Cellular Component (CC)

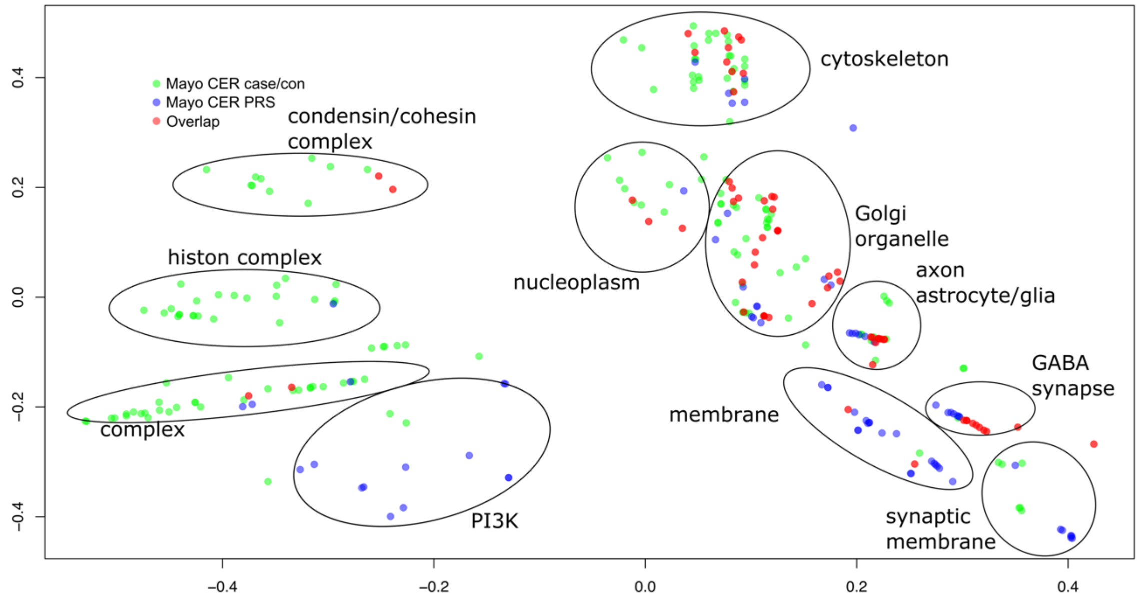


X and Y axes represent CMD dimension 1 and 2. GO term p≤0.05 FDR. Green dots represent significant GO terms from the PRS analysis of cerebellum, Blue dots represent significant GO terms from the PRS analysis of temporal cortex, Red dots represent significant GO terms overlapping in PRS analysis of cerebellum and temporal cortex. Cluster labels were manually curated based on the most common GO term in the cluster.

**Suppl. Figure 21. GO term semantic similarity clustering, cerebellum case/control and PRS (gene order most down-regulated at top; GO down-regulated)**

a) Biological Process (BP)

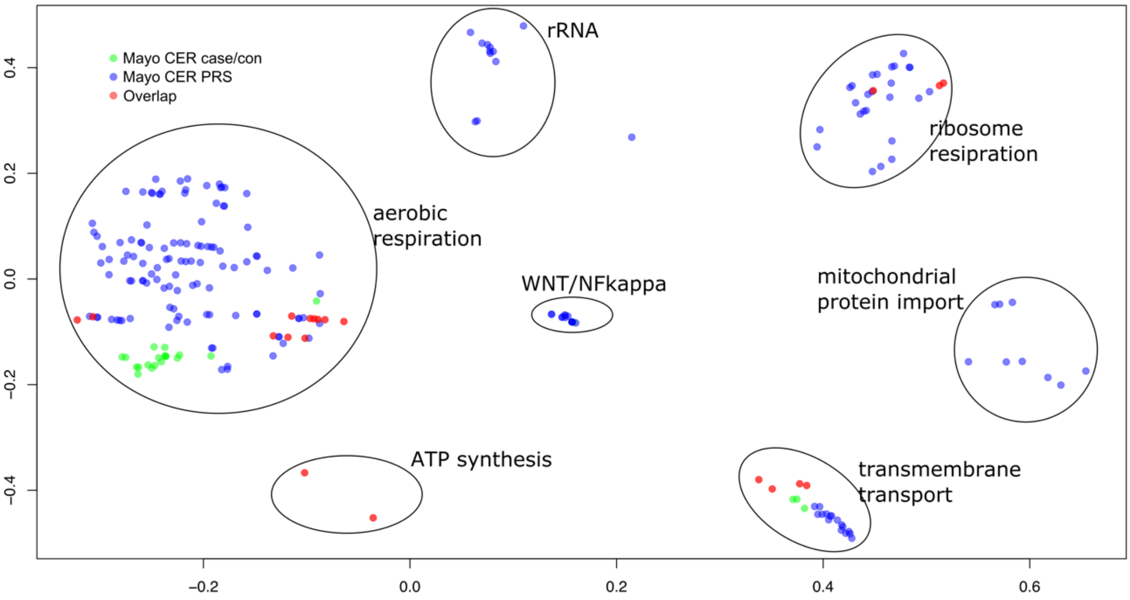


b) Cellular Component (CC)

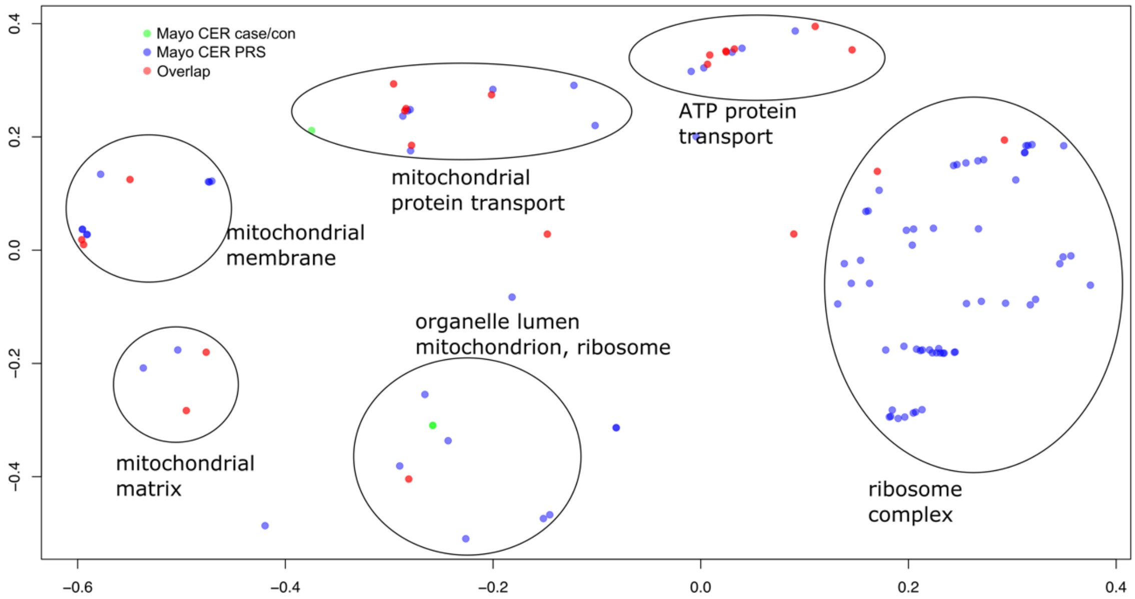


X and Y axes represent CMD dimension 1 and 2. GO term p≤0.05 FDR. Green dots represent significant GO terms from the PRS analysis of cerebellum, Blue dots represent significant GO terms from the PRS analysis of temporal cortex, Red dots represent significant GO terms overlapping in PRS analysis of cerebellum and temporal cortex. Cluster labels were manually curated based on the most common GO term in the cluster.

**Suppl. Figure 22. GO term semantic similarity clustering, temporal cortex case/control and PRS (gene order based on p-values only; GO no direction)**

a) Biological Process (BP)

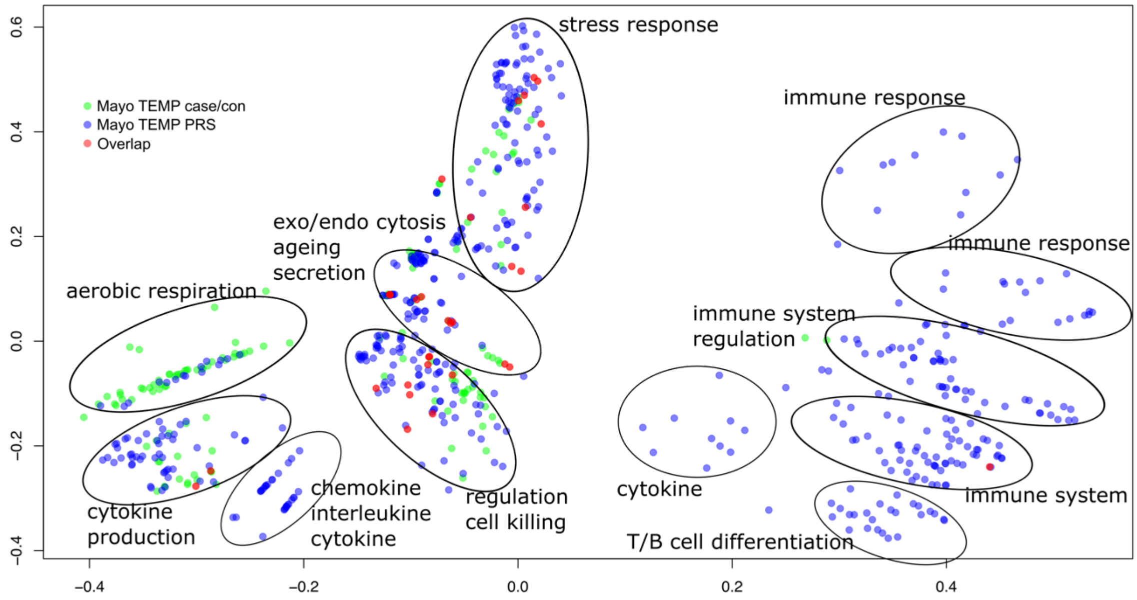


b) Cellular Component (CC)

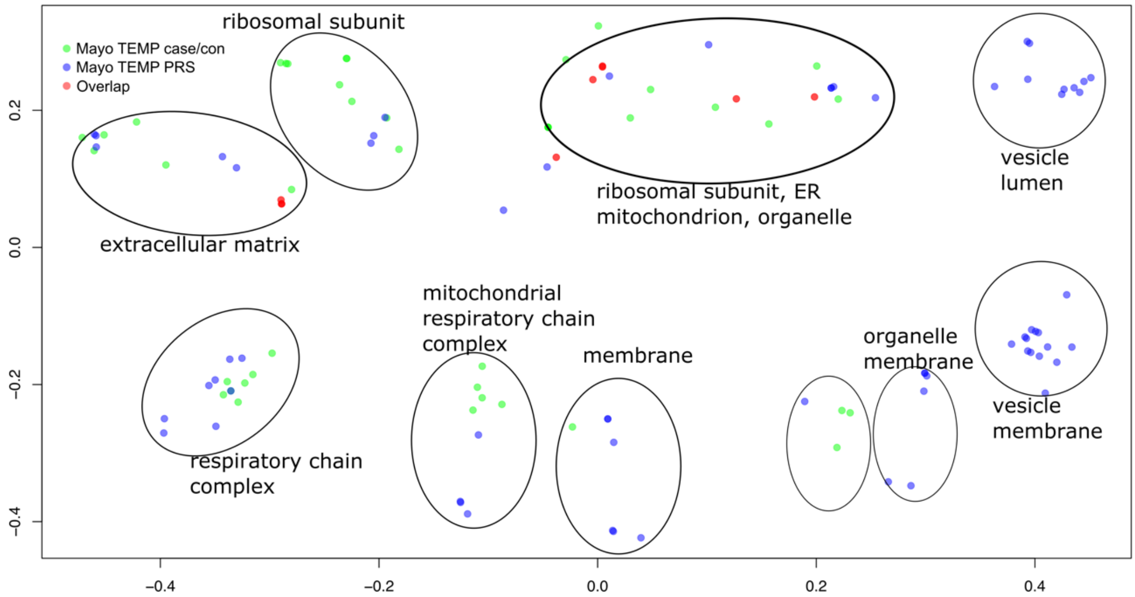


X and Y axes represent CMD dimension 1 and 2. GO term p≤0.05 FDR. Green dots represent significant GO terms from the PRS analysis of cerebellum, Blue dots represent significant GO terms from the PRS analysis of temporal cortex, Red dots represent significant GO terms overlapping in PRS analysis of cerebellum and temporal cortex. Cluster labels were manually curated based on the most common GO term in the cluster.

**Suppl. Figure 23. GO term semantic similarity clustering, temporal cortex case/control and PRS (gene order most up-regulated at top; GO up-regulated)**

a) Biological Process (BP)

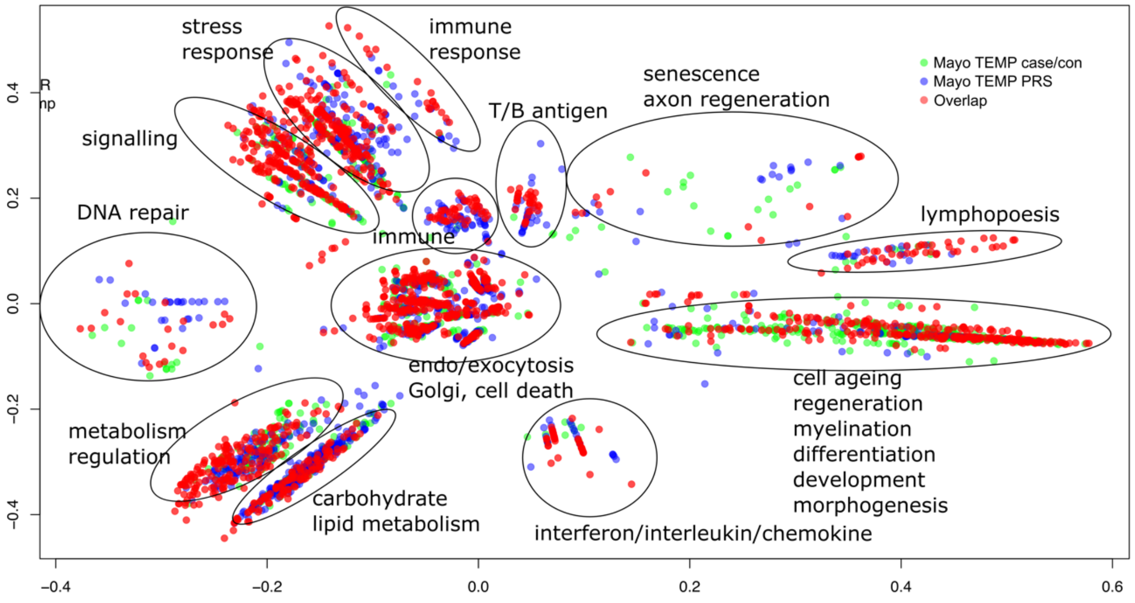


b) Cellular Component (CC)

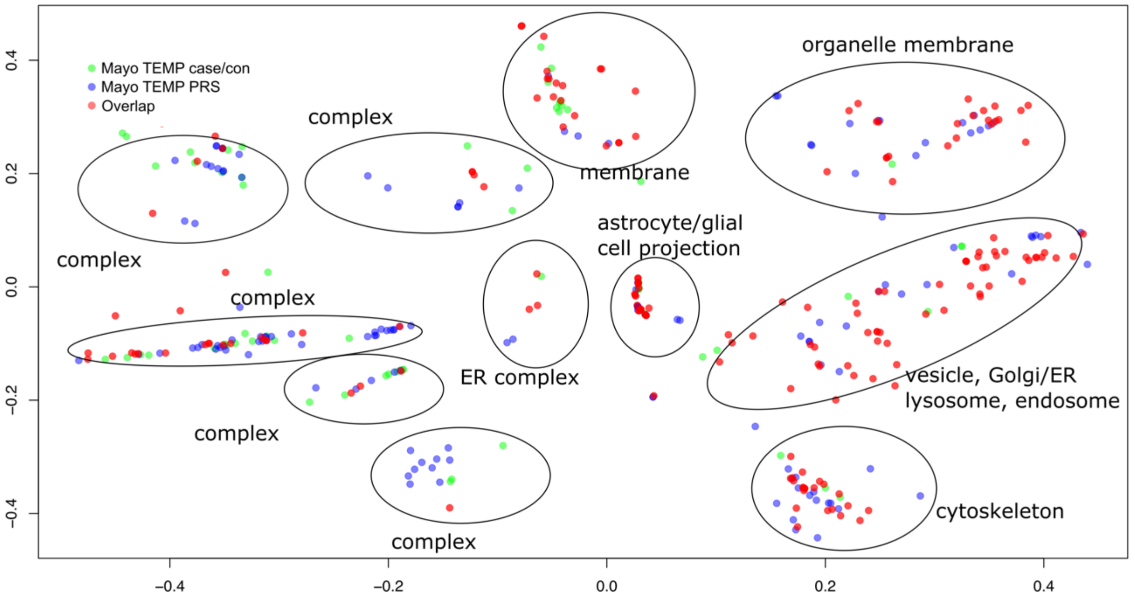


X and Y axes represent CMD dimension 1 and 2. GO term p≤0.05 FDR. Green dots represent significant GO terms from the PRS analysis of cerebellum, Blue dots represent significant GO terms from the PRS analysis of temporal cortex, Red dots represent significant GO terms overlapping in PRS analysis of cerebellum and temporal cortex. Cluster labels were manually curated based on the most common GO term in the cluster.

**Suppl. Figure 24. GO term semantic similarity clustering, temporal cortex case/control and PRS (gene order most down-regulated at top; GO down-regulated)**

a) Biological Process (BP)

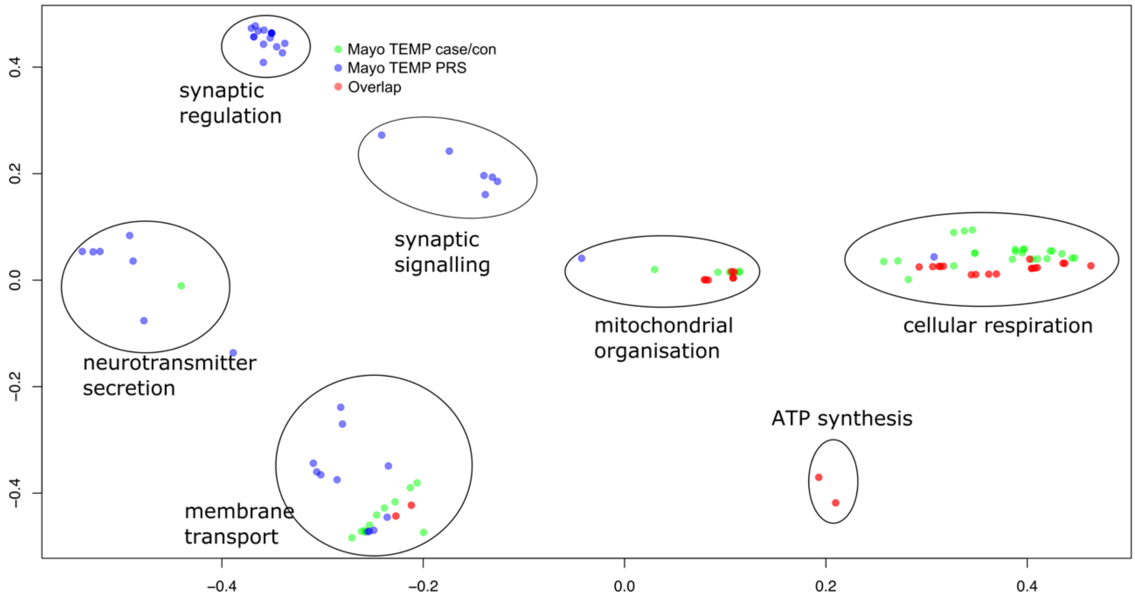


b) Cellular Component (CC)

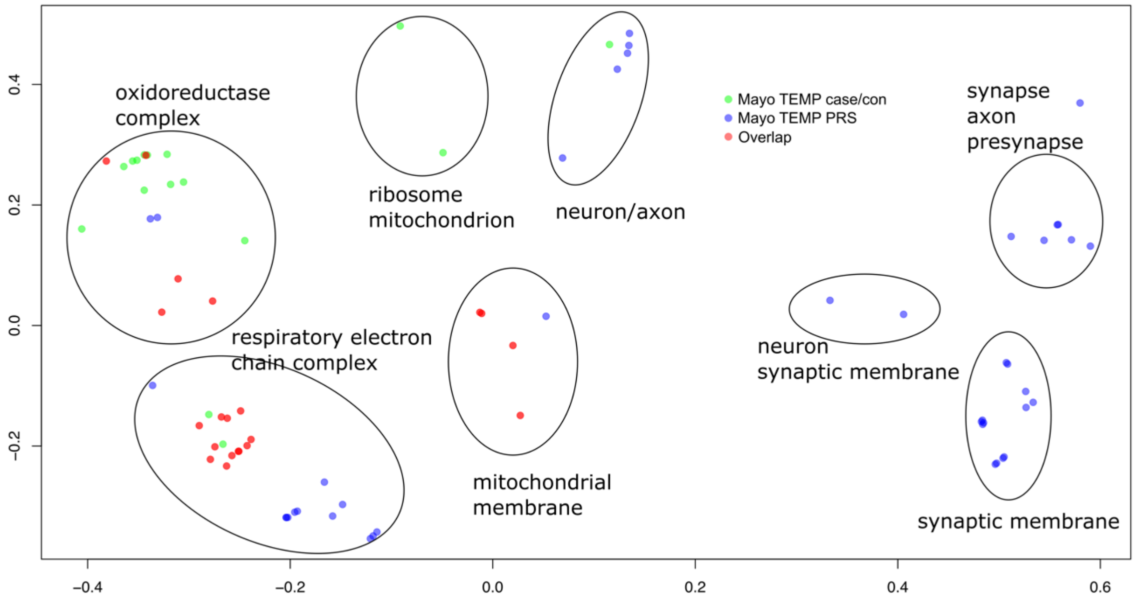


X and Y axes represent CMD dimension 1 and 2. GO term p≤0.05 FDR. Green dots represent significant GO terms from the PRS analysis of cerebellum, Blue dots represent significant GO terms from the PRS analysis of temporal cortex, Red dots represent significant GO terms overlapping in PRS analysis of cerebellum and temporal cortex. Cluster labels were manually curated based on the most common GO term in the cluster.

### 1. Supplementary Materials and Methods

#### 1.1. Overlap with AD disease risk genes

Genes that have been shown to be associated with AD were derived from the largest to-date AD GWAS results (Marioni *et al*. 2018[1]; Kunkle *et al*. 2019[2]; Jansen *et al*. 2019[3]; Lambert *et al*. 2013[4]; Wightman *et al*. 2021[5]; Bellenguez *et al*. 2022[6]). For simplicity, the closest genes to genome-wide significant SNPs were chosen as AD GWAS genes. It is beyond the scope of the work presented here to define the most likely GWAS AD genes. The list of genes is provided in Supplementary Data 3.
One-sided Wilcox-rank sum test was used to determine the statistical significance of AD GWAS genes among the gene-expression results. The gene-expression ranks of the AD GWAS genes were used for the Wilcox-rank sum test.

#### 1.2. Gene Ontology enrichment analysis

The Wilcoxon rank sum test, as implemented in Catmap[7], was used to perform functional analysis, that is significant enrichment of Gene Ontology (GO) categories. Ensembl gene identifiers were mapped to Gene Ontology identifiers (Ensembl Biomart version GRCh38.p13, May 2021) including Molecular Function (MF), Biological Process (BP) and Cellular Component (CC). GO terms relationship was obtained using custom programs based on the go-basic.obo file from http://geneontology.org/ (data-version release 2021-02-01). Ranks of genes were based on the p-value from *DESeq2*. To account for multiple hypotheses testing the Benjamini-Hochberg false discovery rate was used (FDR). Gene lists were sorted by log-fold change (*DESeq2*) and p-value. For all tests three sets of lists were derived; a gene list comprising differentially expressed genes based on p-value only (termed no-direction), a gene list comprising most differentially up-regulated (log-fold change or $\beta > 0$) genes at the top of the list and most differentially down-regulated genes (log-fold change or $\beta < 0$) at the bottom of the list (termed up-to-down) and vice versa (termed down-to-up). If a GO category was found to be statistically significant in the up-to-down list, this GO was referred to as up-regulated, i.e. a large enough proportion of the genes that are part of this GO category were found to be up-regulated or at the top of the list. If a GO category was found to be statistically significant in the down-to-up list, this GO was referred to as down-regulated, i.e. a large enough proportion of the genes that are part of this GO category were found to be down-regulated or at the top of the list.

### 2. References

1. Marioni RE, Harris SE, Zhang Q, McRae AF, Hagenaars SP, Hill WD, et al. GWAS on family history of Alzheimer's disease. Transl Psychiatry. 2018;8:99.
2. Kunkle BW, Grenier-Boley B, Sims R, Bis JC, Damotte V, Naj AC, et al. Genetic meta-analysis of diagnosed Alzheimer's disease identifies new risk loci and implicates Abeta, tau, immunity and lipid processing. Nat Genet. 2019;51:414-30.
3. Jansen IE, Savage JE, Watanabe K, Bryois J, Williams DM, Steinberg S, et al. Genome-wide meta-analysis identifies new loci and functional pathways influencing Alzheimer's disease risk. Nat Genet. 2019;51:404-13.
4. Lambert JC, Ibrahim-Verbaas CA, Harold D, Naj AC, Sims R, Bellenguez C, et al. Meta-analysis of 74,046 individuals identifies 11 new susceptibility loci for Alzheimer's disease. Nat Genet. 2013;45:1452-8.

5. Wightman DP, Jansen IE, Savage JE, Shadrin AA, Bahrami S, Holland D, et al. A genome-wide association study with 1,126,563 individuals identifies new risk loci for Alzheimer's disease. Nat Genet. 2021;53:1276-82.
6. Bellenguez C, Kucukali F, Jansen IE, Kleineidam L, Moreno-Grau S, Amin N, et al. New insights into the genetic etiology of Alzheimer's disease and related dementias. Nat Genet. 2022.
7. Breslin T, Eden P, Krogh M. Comparing functional annotation analyses with Catmap. BMC Bioinformatics. 2004;5:193.
